# Correlates of Daytime Sleepiness and Insomnia among Adults in Samoa

**DOI:** 10.1101/2022.05.25.22275570

**Authors:** Lacey W. Heinsberg, Jenna C. Carlson, Alysa Pomer, Brian E. Cade, Take Naseri, Muagututia Sefuiva Reupena, Daniel E. Weeks, Stephen T. McGarvey, Susan Redline, Nicola L. Hawley

## Abstract

**Objective:** To describe daytime sleepiness and insomnia among adults in Samoa and identify modifiable factors associated with these measures.

**Design/setting:** Cross-sectional analysis of data from the *Soifua Manuia* (“Good Health”) study (n=519, 55.1% female); Upolu island, Samoa.

**Methods:** Daytime sleepiness and insomnia were assessed with the Epworth Sleepiness Scale (ESS) and the Women’s Health Initiative Insomnia Rating Scale (WHIIRS), respectively. Detailed physical, sociodemographic, and behavioral factors were collected. Sleep measures were characterized using multiple linear regression with backwards elimination and a bootstrap stability investigation.

**Results:** Excessive daytime sleepiness (ESS>10) and insomnia (WHIIRS>10) were reported by 20% and 6.3% of the sample, respectively. ESS scores were higher in individuals reporting more physical activity (Estimate=1.88; 95% CI=1.12 to 2.75), higher material wealth (0.18; 0.09 to 0.28), and asthma (2.85; 1.25 to 4.51). ESS scores were lower in individuals residing in periurban versus urban regions (−1.43; −2.39 to −0.41), reporting no work versus day shift work (−2.26; −3.07 to −1.41), and reporting greater perceived stress (−0.14; −0.23 to −0.06). WHIIRS scores were lower in individuals reporting “other” shift work (split/irregular/on-call/rotating) versus day shift work (−1.96; −2.89 to −1.14) and those who perceived their village’s wealth to be poor/average versus wealthy (−0.94; −1.50 to −0.34).

**Conclusions:** Participants had a generally higher prevalence of excessive daytime sleepiness, but lower prevalence of insomnia, compared with individuals from high-income countries. Factors associated with sleep health differed compared with prior studies, emphasizing potential cultural/environmental differences and the need for targeted interventions to improve sleep health in this setting.

## 1. INTRODUCTION

Daytime sleepiness and sleep quality, including insomnia symptoms, are important public health issues given their associations with traffic/workplace accidents, cardiometabolic diseases, and mortality [1–3]. There are several social factors associated with sleep health, including those indexed by race/ethnicity [1, 4]. Notably, Pacific Islanders in high-income countries experience a high prevalence of sleep disturbances [5–7]. For example, as part of the Native Hawaiian and Pacific Islander National Health Interview Survey, individuals in the United States (US) who self-identified as Native Hawaiian or Pacific Islander reported shorter sleep duration and poorer sleep quality than individuals who self-identified as White [7]. Both were independently associated with hypertension and diabetes, highlighting the importance of sleep for health and wellness among this population [7]. Likewise, in New Zealand, Maori and Pacific peoples were particularly likely to report short sleep duration (<7 hours), which was negatively associated with psychological well-being regardless of health and personality factors [8].

Unfortunately, little high-quality sleep data from individuals residing in low- and middle-income countries exists [9], including the Pacific Island nation of Samoa. Sleep environments in low- and middle-income countries differ from high-income countries in ways that may positively or negatively impact sleep health. In Samoa, (1) exposure to heat/humidity/rain in traditional open-air *fales* (homes), (2) time-consuming work-related travel if an individual’s residence is beyond the single urban center, (3) differences in ambient lighting across geographic regions (i.e., rural versus urban), (4) large, multigenerational families (many people sharing rooms/beds for sleeping), and (5) sleeping on *fale moe* (palm mats) may all contribute to a sleep health risk profile that differs from high-income populations.

Potentially contributing to poor sleep quality and daytime sleepiness, Samoa has the 8^th^ highest percentage of adult obesity in the world [10]; in 2013 (the most recent data available) 53.1% of men and 76.7% of women had obesity based on a BMI ≥30 kg/m^2^ [11]. This is attributable in large part to rapid economic and nutritional transition characterized by increased access to micronutrient-poor, energy-dense foods and a decrease in subsistence-related physical activity [12]. Obesity has been shown to play a role in sleep health in high-income countries [13, 14], but it is unclear if this association exists in low- and middle-income countries such as Samoa where obesity prevalence is high. Therefore, the purpose of this study was to provide the first-ever description of daytime sleepiness and insomnia among adults from Samoa, including the identification of potentially modifiable factors associated with these measures.

## 2. MATERIALS AND METHODS

### 2.1 Study design, setting, and sample

This was a secondary analysis of data collected as part of the 2017-2019 *Soifua Manuia* (“Good Health”) study, which aimed to characterize the impact of a genetic variant (rs373863828, c.1370G>A, p.R457Q) in CREB3 Regulatory Factor (*CREBRF*) on metabolic and behavioral traits in adults from Samoa [15–17]. The study protocol is described in detail elsewhere [17]. In brief, participants were purposively recruited based on their earlier participation in a 2010 genome-wide association study [18]. Recruitment procedures included intentional oversampling of the *CREBRF* obesity-risk allele (A) [17, 18]. Participants from the island of ‘Upolu, residing in the Apia Urban Area (AUA, urban), Northwest ‘Upolu (NWU, periurban), and Rest of ‘Upolu (ROU, rural) census regions were included. Exclusion criteria included (1) pregnancy, giving birth within 6 months, or current lactation, (2) physical or cognitive impairment preventing full participation, or (3) use of weight loss medications, history of weight loss surgery, or weight loss of >5% of body weight in the past year. Research assistants attempted to contact 770 participants, and screened 555; 519 (n=286 female) were eligible and participated in data collection [17]. The study was approved by Institutional Review Boards at Yale University, Brown University, the University of Pittsburgh, Mass General Brigham (formerly Partners Healthcare), and the Health Research Committee of the Samoa Ministry of Health. All participants gave written informed consent.

### 2.2 Participant characteristics

Participants completed extensive physical assessments and comprehensive questionnaires administered by trained, bilingual Samoan research assistants [17]. Variables used in this analysis included those significantly associated with insomnia or daytime sleepiness in the literature (e.g., work schedule [19]) and those unique to the Samoan population (e.g., number of household assets, community spirit, census region of residence).

#### 2.2.1 Biological factors and health history

Age and sex were self-reported. Weight and height were measured in duplicate to the nearest 0.1 kg and 0.1 cm using a digital weighing scale (Tanita HD 351; Tanita Corporation of America) and portable stadiometer (SECA 213, Seca GmbH & Co), averaged, and used to compute BMI. Abdominal circumference was measured in duplicate to the nearest 0.1 cm at the level of the umbilicus and averaged. *CREBRF* rs373863828 genotype data, generated via TaqMan real-time PCR (Applied Biosystems), came from the earlier 2010 GWAS study [16]. Based on current use of diabetes medication or Hemoglobin A1c (HbA1c) values, participants were categorized as having no diabetes (no diabetes medication and HbA1c <5.7%), pre-diabetes (no diabetes medication and HbA1c of 5.7-6.4%), or type 2 diabetes (diabetes medication and/or HbA1c>6.4%). Blood pressure (BP) was measured in triplicate and the second and third values averaged for analyses. Hypertension was defined as an average BP ≥140/90 mmHg, in line with local diagnostic criteria, or hypertension medication use. Asthma diagnoses were self-reported by participants. The Short Form-8 (SF-8) was used to assess perceived physical and mental health using the Physical Health Component Score and Mental Health Component Score summary measures. Both result in values of 0-100 with higher scores indicating better perceived health [20].

#### 2.2.2 Sociodemographic and behavioral factors

Census region of residence was based on the three geographic regions described above (AUA/NWU/ROU). A material lifestyle score, a proxy for socioeconomic resources used extensively in Samoa, was computed as the sum of items owned from an 18-item household inventory (refrigerator, freezer, portable stereo/MP3 player, etc.) [17]. Additional self-reported sociodemographic factors included years of education, relationship status (partnered/not partnered), children at home (yes/no), and work schedule (due to low cell count, categorized as day shift/afternoon; night shift/other [split/irregular/on call/rotating shifts]; or does not work). The Global Physical Activity Questionnaire was used to estimate self-reported daily moderate-to-vigorous physical activity (MVPA) minutes [21] (dichotomized as 0/>0 MVPA minutes based on an extreme floor effect seen in this population).

Additional social data of interest included home ownership (own/rent), smoking (current use of cigarettes, tobacco, or pipe; yes/no), and consumption of alcohol in the last 12 months (yes/no). Participants also reported their perception of the wealth and community spirit of their village of residence (poor-average/wealthy and low-average/strong, respectively). Psychosocial variables included perceived stress, social conflict, and social support. Perceived stress was measured using Cohen’s Perceived Stress Scale (scores 0 to 40) [22] and social conflict was measured using the Perceived Social Conflict Score (scores from 6 to 30) [23]; higher scores indicated higher perceived stress and social conflict, respectively. Finally, the total score of the Multidimensional Scale of Perceived Social Support (MSPSS, scores 1 to 5), which includes questions related to support from family, friends, and an individual’s significant other, was used to measure social support, with higher scores indicating more perceived support [24]. Additional details of the questionnaires and variables of interest are provided in Supplementary Information (SI, Table S1).

#### 2.2.3 Sleep measures

Primary sleep outcomes (i.e., dependent variables) included self-reported daytime sleepiness and insomnia symptoms (sleep quality) measured using the Epworth Sleepiness Scale (ESS) [25] and the Women’s Health Initiative Insomnia Rating Scale (WHIIRS) [26]. The ESS and WHIIRS range from 0 to 24 and 0 to 20, respectively, with higher scores indicating greater severity of daytime sleepiness or insomnia. Though these tools had not been used in Samoa previously, the ESS has an alpha coefficient of 0.88 and test-retest reliability coefficient of 0.82 (5 months apart) [27]. Likewise, the WHIIRS is valid in a variety of populations with an alpha coefficient of 0.78 and test-retest reliability coefficients of 0.96 (same-day administration) and 0.66 (≥1 year apart) [26].

Additional sleep measures of interest included categorical excessive daytime sleepiness and insomnia (yes/no) calculated using ESS and WHIIRS cut points of >10 as applied elsewhere [4]; self-reported chronotype (morning person, evening person); self-reported use of sleeping pills in the past four weeks (yes/no); and self-reported sleep duration per weeknight or weekday (depending on work schedule) both as a continuous variable, and categorized as short sleep (<6 hours [4]), sufficient sleep (6-9 hours), or long sleep (>9 hours [28]).

### 2.3 Statistical analyses

All statistical analyses were performed in R version 3.6.0. Standard descriptive statistics were computed including means, standard deviations (SD), medians, and minimum/maximum values for continuous variables and frequency counts and percentages for categorical variables. Data were examined graphically and statistically to identify outliers and assess patterns of missing data. Within-domain imputation was performed using the mice package [29] in R for continuous summary score variables with a missingness of 5-10% (i.e., SF-8 Physical and Mental Health, material lifestyle score, Cohen’s Perceived Stress Scale, the MSPSS, Perceived Social Conflict Score, ESS, and WHIIRS) as detailed in the SI (Table S2), and sensitivity analyses were performed to assess the influence of imputation.

Preliminary analyses were performed to evaluate bivariate associations between participant factors and sleep outcomes (ESS and WHIIRS). Depending on variable type and distribution, relationships were evaluated using Pearson or Spearman correlations, t-tests, or one-way analysis of variance (ANOVA). Where the assumption of constant variance was violated, Welch’s one-way analysis of means was used to evaluate relationships between groups. Given sex-specific health differences in the Samoan population [11, 15], and the role of sex in sleep health [30], bivariate analyses were performed both for the overall sample and stratified by sex.

As the first study to examine sleep health in the Samoan population, and without strong theory to support *a priori* modeling, multiple linear regression and backwards elimination with Akaike information criterion (AIC) was used for variable selection [31], treating ESS or WHIIRS as the outcome variables. Based on the results of bivariate analyses (suggesting few ESS-or WHIIRS-related sex differences in this sample as summarized below), sex stratification was not carried forward for the multivariable modeling so that the largest sample size could be retained. However, sex was included as a covariate in all regression models along with age, BMI, census region, and rs373863828 genotype (additively coded based on number of minor allele (A) copies) based on *a priori* knowledge of the Samoan population or to adjust for study design. Additional variables of interest included those described above. Only individuals with complete data post-imputation for all 28 variables of interest could be included in the multivariable modeling (n=443), making the global model number of events per variable (EPV_global_) 443/28=15.8. Given the potential for variable-selection bias and issues associated with the use of the same data set for both variable selection and post-selection inference, a bootstrap stability investigation was performed to both assess the impact of variable selection and perform multi-model inference as recommended by Heinze, Wallisch, and Dunkler [31]. As part of this process, the bootstrap-inclusion frequency was calculated for each variable of interest and variation in regression coefficients around the full model estimates was assessed using bootstrapped resamples, repeating the backwards elimination selection procedure 1,000 times.

Coefficients and 95% confidence intervals (CI) were reported for the global model (including all input variables of interest) and the selected model (including variables chosen by backwards elimination). Final multi-model coefficients and 95% CI used for inference were reported as the median of the bootstrap distribution with multiple comparison interpretation built into the modeling approach. The root mean squared difference (RMSD) ratio, relative conditional bias, and model selection frequencies were calculated to further assess variable-selection bias as described in the SI. Selected model assessment was performed using residual analysis, collinearity, and influence diagnostics. To stabilize regression parameters, variables were collapsed as described above and further detailed in the SI (Table S1).

## 3. RESULTS

### 3.1 Sample characteristics and unadjusted analyses

The overall sample consisted of 519 participants (55.1% female) with a mean (±SD) age of 52.2 (±10.0) years and BMI of 35.8 (±7.7) kg/m^2^ (Table 1). ESS scores ranged from 0 to 16 with a mean score of 6.9 (±4.1) with 20.0% of individuals meeting the threshold for excessive daytime sleepiness (ESS >10 [4]). WHIIRS scores ranged from 0 to 16 with a mean score of 6.5 (±2.7) with 6.3% of individuals having a WHIIRS score consistent with clinical thresholds for insomnia (WHIIRS >10 [4]).

**Table 1.**
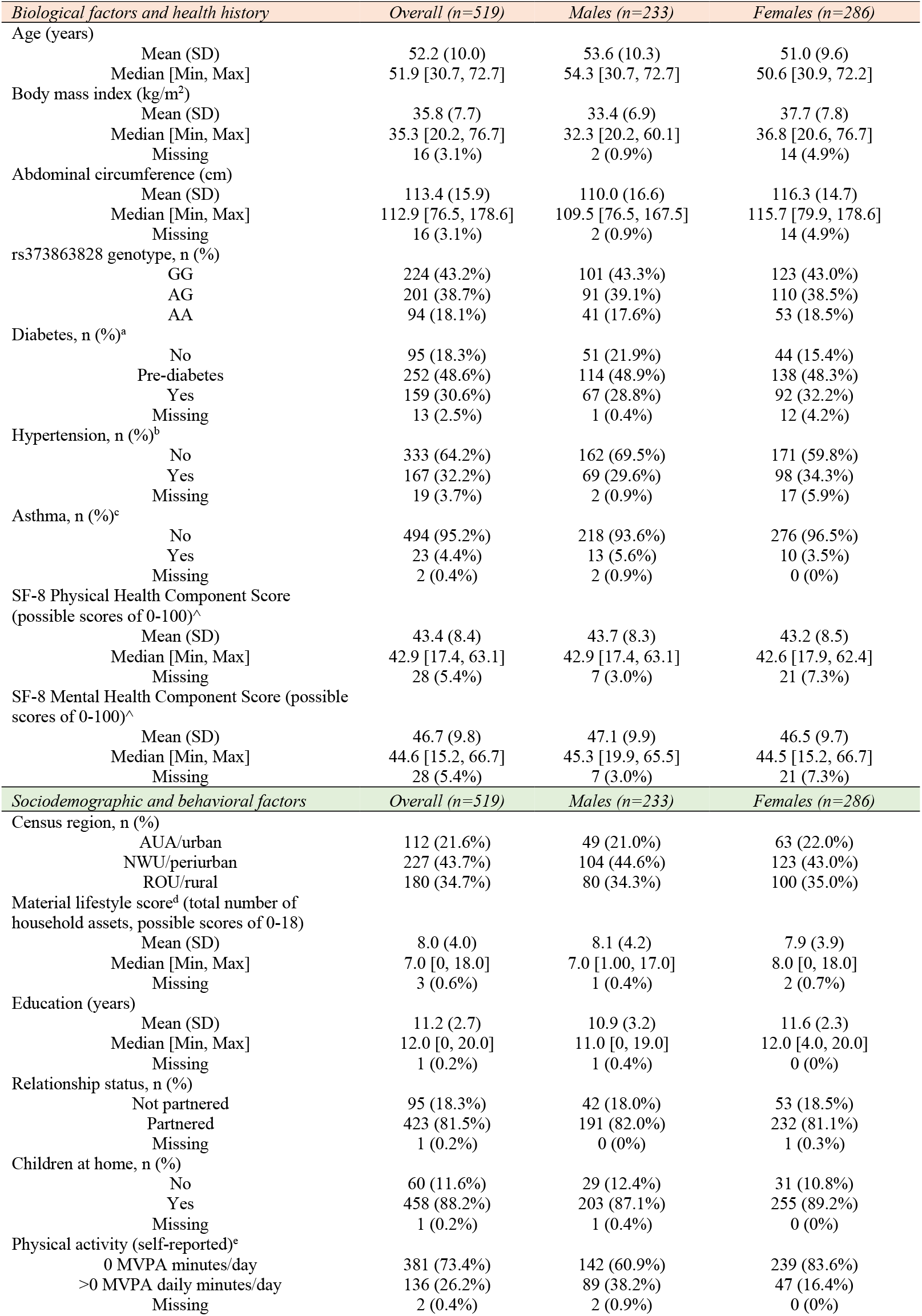
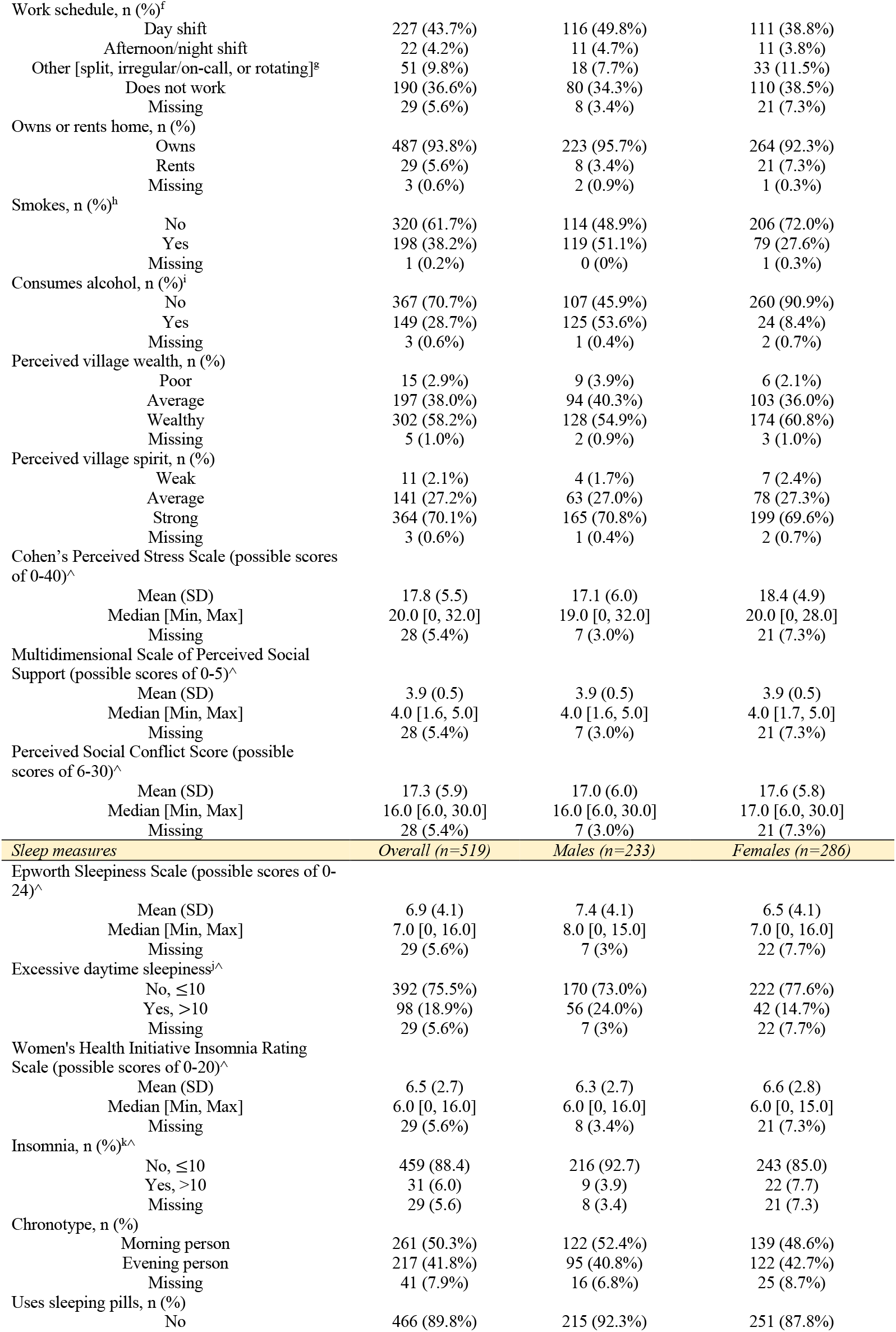
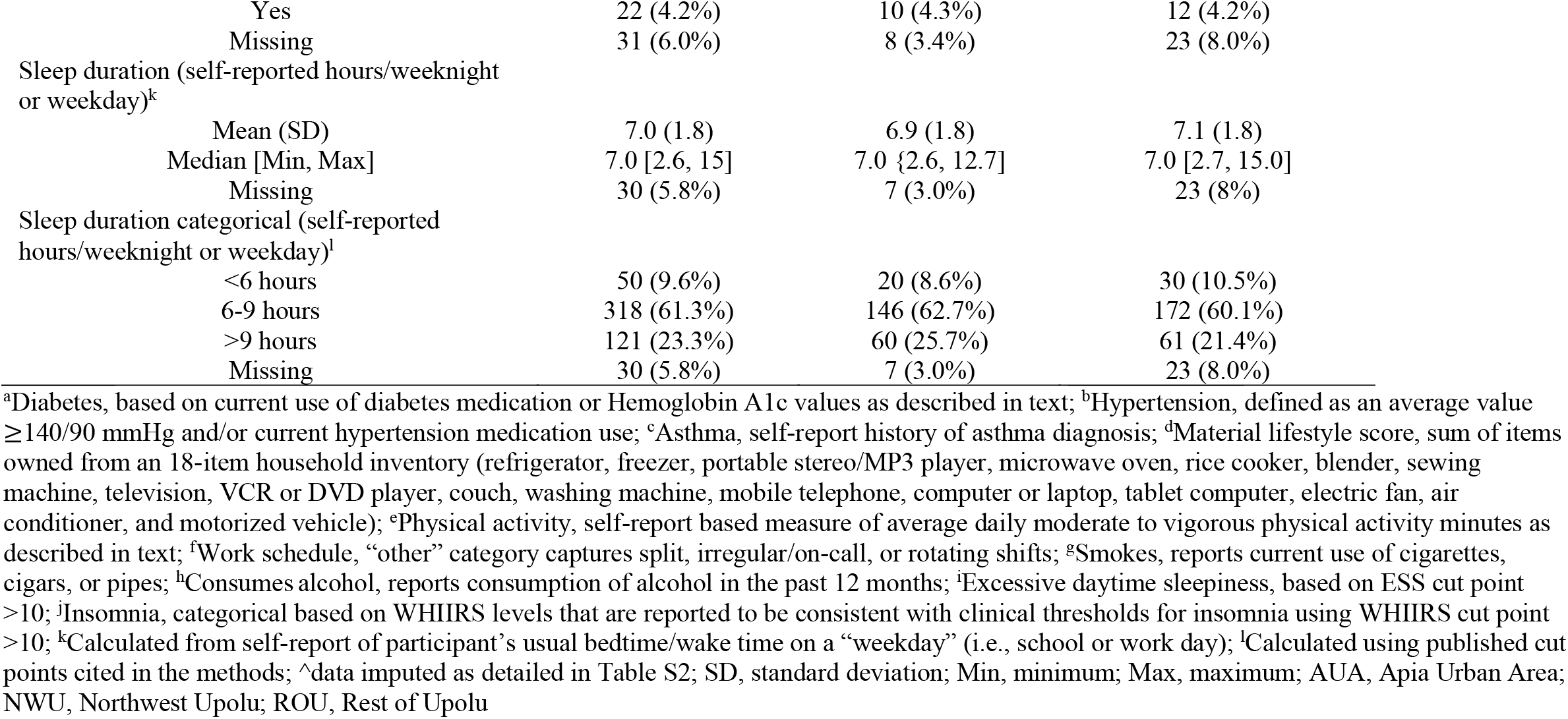
Sample characteristics

| *Biological factors and health history* | *Overall (n=519)* | *Males (n=233)* | *Females (n=286)* |
| --- | --- | --- | --- |
| Age (years) |  |  |  |
| Mean (SD) | 52.2 (10.0) | 53.6 (10.3) | 51.0 (9.6) |
| Median [Min, Max] | 51.9 [30.7, 72.7] | 54.3 [30.7, 72.7] | 50.6 [30.9, 72.2] |
| Body mass index (kg/m<sup>2</sup>) |  |  |  |
| Mean (SD) | 35.8 (7.7) | 33.4 (6.9) | 37.7 (7.8) |
| Median [Min, Max] | 35.3 [20.2, 76.7] | 32.3 [20.2, 60.1] | 36.8 [20.6, 76.7] |
| Missing | 16 (3.1%) | 2 (0.9%) | 14 (4.9%) |
| Abdominal circumference (cm) |  |  |  |
| Mean (SD) | 113.4 (15.9) | 110.0 (16.6) | 116.3 (14.7) |
| Median [Min, Max] | 112.9 [76.5, 178.6] | 109.5 [76.5, 167.5] | 115.7 [79.9, 178.6] |
| Missing | 16 (3.1%) | 2 (0.9%) | 14 (4.9%) |
| rs373863828 genotype, n (%) |  |  |  |
| GG | 224 (43.2%) | 101 (43.3%) | 123 (43.0%) |
| AG | 201 (38.7%) | 91 (39.1%) | 110 (38.5%) |
| AA | 94 (18.1%) | 41 (17.6%) | 53 (18.5%) |
| Diabetes, n (%)<sup>a</sup> |  |  |  |
| No | 95 (18.3%) | 51 (21.9%) | 44 (15.4%) |
| Pre-diabetes | 252 (48.6%) | 114 (48.9%) | 138 (48.3%) |
| Yes | 159 (30.6%) | 67 (28.8%) | 92 (32.2%) |
| Missing | 13 (2.5%) | 1 (0.4%) | 12 (4.2%) |
| Hypertension, n (%)<sup>b</sup> |  |  |  |
| No | 333 (64.2%) | 162 (69.5%) | 171 (59.8%) |
| Yes | 167 (32.2%) | 69 (29.6%) | 98 (34.3%) |
| Missing | 19 (3.7%) | 2 (0.9%) | 17 (5.9%) |
| Asthma, n (%)<sup>c</sup> |  |  |  |
| No | 494 (95.2%) | 218 (93.6%) | 276 (96.5%) |
| Yes | 23 (4.4%) | 13 (5.6%) | 10 (3.5%) |
| Missing | 2 (0.4%) | 2 (0.9%) | 0 (0%) |
| SF-8 Physical Health Component Score<br>(possible scores of 0-100)<sup>^</sup> |  |  |  |
| Mean (SD) | 43.4 (8.4) | 43.7 (8.3) | 43.2 (8.5) |
| Median [Min, Max] | 42.9 [17.4, 63.1] | 42.9 [17.4, 63.1] | 42.6 [17.9, 62.4] |
| Missing | 28 (5.4%) | 7 (3.0%) | 21 (7.3%) |
| SF-8 Mental Health Component Score (possible<br>scores of 0-100)<sup>^</sup> |  |  |  |
| Mean (SD) | 46.7 (9.8) | 47.1 (9.9) | 46.5 (9.7) |
| Median [Min, Max] | 44.6 [15.2, 66.7] | 45.3 [19.9, 65.5] | 44.5 [15.2, 66.7] |
| Missing | 28 (5.4%) | 7 (3.0%) | 21 (7.3%) |
| *Sociodemographic and behavioral factors* | *Overall (n=519)* | *Males (n=233)* | *Females (n=286)* |
| Census region, n (%) |  |  |  |
| AUA/urban | 112 (21.6%) | 49 (21.0%) | 63 (22.0%) |
| NWU/periurban | 227 (43.7%) | 104 (44.6%) | 123 (43.0%) |
| ROU/rural | 180 (34.7%) | 80 (34.3%) | 100 (35.0%) |
| Material lifestyle score<sup>d</sup> (total number of<br>household assets, possible scores of 0-18) |  |  |  |
| Mean (SD) | 8.0 (4.0) | 8.1 (4.2) | 7.9 (3.9) |
| Median [Min, Max] | 7.0 [0, 18.0] | 7.0 [1.00, 17.0] | 8.0 [0, 18.0] |
| Missing | 3 (0.6%) | 1 (0.4%) | 2 (0.7%) |
| Education (years) |  |  |  |
| Mean (SD) | 11.2 (2.7) | 10.9 (3.2) | 11.6 (2.3) |
| Median [Min, Max] | 12.0 [0, 20.0] | 11.0 [0, 19.0] | 12.0 [4.0, 20.0] |
| Missing | 1 (0.2%) | 1 (0.4%) | 0 (0%) |
| Relationship status, n (%) |  |  |  |
| Not partnered | 95 (18.3%) | 42 (18.0%) | 53 (18.5%) |
| Partnered | 423 (81.5%) | 191 (82.0%) | 232 (81.1%) |
| Missing | 1 (0.2%) | 0 (0%) | 1 (0.3%) |
| Children at home, n (%) |  |  |  |
| No | 60 (11.6%) | 29 (12.4%) | 31 (10.8%) |
| Yes | 458 (88.2%) | 203 (87.1%) | 255 (89.2%) |
| Missing | 1 (0.2%) | 1 (0.4%) | 0 (0%) |
| Physical activity (self-reported)<sup>e</sup> |  |  |  |
| 0 MVPA minutes/day | 381 (73.4%) | 142 (60.9%) | 239 (83.6%) |
| >0 MVPA daily minutes/day | 136 (26.2%) | 89 (38.2%) | 47 (16.4%) |
| Missing | 2 (0.4%) | 2 (0.9%) | 0 (0%) |
| Work schedule, n (%)<sup>f</sup> |  |  |  |
| Day shift | 227 (43.7%) | 116 (49.8%) | 111 (38.8%) |
| Afternoon/night shift | 22 (4.2%) | 11 (4.7%) | 11 (3.8%) |
| Other [split, irregular/on-call, or rotating]<sup>g</sup> | 51 (9.8%) | 18 (7.7%) | 33 (11.5%) |
| Does not work | 190 (36.6%) | 80 (34.3%) | 110 (38.5%) |
| Missing | 29 (5.6%) | 8 (3.4%) | 21 (7.3%) |
| Owens or rents home, n (%) |  |  |  |
| Owns | 487 (93.8%) | 223 (95.7%) | 264 (92.3%) |
| Rents | 29 (5.6%) | 8 (3.4%) | 21 (7.3%) |
| Missing | 3 (0.6%) | 2 (0.9%) | 1 (0.3%) |
| Smokes, n (%)<sup>h</sup> |  |  |  |
| No | 320 (61.7%) | 114 (48.9%) | 206 (72.0%) |
| Yes | 198 (38.2%) | 119 (51.1%) | 79 (27.6%) |
| Missing | 1 (0.2%) | 0 (0%) | 1 (0.3%) |
| Consumes alcohol, n (%)<sup>i</sup> |  |  |  |
| No | 367 (70.7%) | 107 (45.9%) | 260 (90.9%) |
| Yes | 149 (28.7%) | 125 (53.6%) | 24 (8.4%) |
| Missing | 3 (0.6%) | 1 (0.4%) | 2 (0.7%) |
| Perceived village wealth, n (%) |  |  |  |
| Poor | 15 (2.9%) | 9 (3.9%) | 6 (2.1%) |
| Average | 197 (38.0%) | 94 (40.3%) | 103 (36.0%) |
| Wealthy | 302 (58.2%) | 128 (54.9%) | 174 (60.8%) |
| Missing | 5 (1.0%) | 2 (0.9%) | 3 (1.0%) |
| Perceived village spirit, n (%) |  |  |  |
| Weak | 11 (2.1%) | 4 (1.7%) | 7 (2.4%) |
| Average | 141 (27.2%) | 63 (27.0%) | 78 (27.3%) |
| Strong | 364 (70.1%) | 165 (70.8%) | 199 (69.6%) |
| Missing | 3 (0.6%) | 1 (0.4%) | 2 (0.7%) |
| Cohen's Perceived Stress Scale (possible scores of 0-40)<sup>^</sup> |  |  |  |
| Mean (SD) | 17.8 (5.5) | 17.1 (6.0) | 18.4 (4.9) |
| Median [Min, Max] | 20.0 [0, 32.0] | 19.0 [0, 32.0] | 20.0 [0, 28.0] |
| Missing | 28 (5.4%) | 7 (3.0%) | 21 (7.3%) |
| Multidimensional Scale of Perceived Social Support (possible scores of 0-5)<sup>^</sup> |  |  |  |
| Mean (SD) | 3.9 (0.5) | 3.9 (0.5) | 3.9 (0.5) |
| Median [Min, Max] | 4.0 [1.6, 5.0] | 4.0 [1.6, 5.0] | 4.0 [1.7, 5.0] |
| Missing | 28 (5.4%) | 7 (3.0%) | 21 (7.3%) |
| Perceived Social Conflict Score (possible scores of 6-30)<sup>^</sup> |  |  |  |
| Mean (SD) | 17.3 (5.9) | 17.0 (6.0) | 17.6 (5.8) |
| Median [Min, Max] | 16.0 [6.0, 30.0] | 16.0 [6.0, 30.0] | 17.0 [6.0, 30.0] |
| Missing | 28 (5.4%) | 7 (3.0%) | 21 (7.3%) |
| *Sleep measures* | *Overall (n=519)* | *Males (n=233)* | *Females (n=286)* |
| Epworth Sleepiness Scale (possible scores of 0-24)<sup>^</sup> |  |  |  |
| Mean (SD) | 6.9 (4.1) | 7.4 (4.1) | 6.5 (4.1) |
| Median [Min, Max] | 7.0 [0, 16.0] | 8.0 [0, 15.0] | 7.0 [0, 16.0] |
| Missing | 29 (5.6%) | 7 (3%) | 22 (7.7%) |
| Excessive daytime sleepiness<sup>i^</sup> |  |  |  |
| No, ≤10 | 392 (75.5%) | 170 (73.0%) | 222 (77.6%) |
| Yes, >10 | 98 (18.9%) | 56 (24.0%) | 42 (14.7%) |
| Missing | 29 (5.6%) | 7 (3%) | 22 (7.7%) |
| Women's Health Initiative Insomnia Rating Scale (possible scores of 0-20)<sup>^</sup> |  |  |  |
| Mean (SD) | 6.5 (2.7) | 6.3 (2.7) | 6.6 (2.8) |
| Median [Min, Max] | 6.0 [0, 16.0] | 6.0 [0, 16.0] | 6.0 [0, 15.0] |
| Missing | 29 (5.6%) | 8 (3.4%) | 21 (7.3%) |
| Insomnia, n (%)<sup>k^</sup> |  |  |  |
| No, ≤10 | 459 (88.4) | 216 (92.7) | 243 (85.0) |
| Yes, >10 | 31 (6.0) | 9 (3.9) | 22 (7.7) |
| Missing | 29 (5.6) | 8 (3.4) | 21 (7.3) |
| Chronotype, n (%) |  |  |  |
| Morning person | 261 (50.3%) | 122 (52.4%) | 139 (48.6%) |
| Evening person | 217 (41.8%) | 95 (40.8%) | 122 (42.7%) |
| Missing | 41 (7.9%) | 16 (6.8%) | 25 (8.7%) |
| Uses sleeping pills, n (%) |  |  |  |
| No | 466 (89.8%) | 215 (92.3%) | 251 (87.8%) |
| Yes | 22 (4.2%) | 10 (4.3%) | 12 (4.2%) |
| Missing | 31 (6.0%) | 8 (3.4%) | 23 (8.0%) |
| Sleep duration (self-reported hours/weeknight or weekday)<sup>k</sup> |  |  |  |
| Mean (SD) | 7.0 (1.8) | 6.9 (1.8) | 7.1 (1.8) |
| Median [Min, Max] | 7.0 [2.6, 15] | 7.0 {2.6, 12.7} | 7.0 [2.7, 15.0] |
| Missing | 30 (5.8%) | 7 (3.0%) | 23 (8%) |
| Sleep duration categorical (self-reported hours/weeknight or weekday)<sup>l</sup> |  |  |  |
| <6 hours | 50 (9.6%) | 20 (8.6%) | 30 (10.5%) |
| 6-9 hours | 318 (61.3%) | 146 (62.7%) | 172 (60.1%) |
| >9 hours | 121 (23.3%) | 60 (25.7%) | 61 (21.4%) |
| Missing | 30 (5.8%) | 7 (3.0%) | 23 (8.0%) |

The results of bivariate analyses examining associations between participant characteristics and sleep outcomes for the overall sample, and stratified by sex, are presented in the SI for ESS (Table S3) and WHIIRS (Table S4). Of note, statistically significant unadjusted differences in daytime sleepiness were observed across several factors. Specifically, ESS scores were: higher in males than females; higher with each copy of the rs373863828 minor allele (A); higher in those with asthma than without; negatively correlated with self-reported physical health; lowest in individuals residing in NWU (periurban); positively correlated with material lifestyle score; higher in individuals reporting >0 MVPA minutes/day; highest in individuals with afternoon/night work schedules and second highest in the “other” shift work (i.e., split/irregular/on call/rotating shifts); higher in individuals who smoked and consumed alcohol than those who did not; higher in individuals who perceive their village spirit to be weak/average; negatively correlated with perceived stress; positively correlated with perceived social support; and higher in individuals who reported being an “evening person”. With the exception of potential sex-specific differences in ESS scores based on SF-8 Physical and Mental health, alcohol consumption, social support, and chronotype, the results were generally concordant between sexes (Table S3).

Based on bivariate analyses, WHIIRS scores were: positively correlated with BMI, abdominal circumference, and years of education; highest in individuals residing in NWU (periurban); lowest in individuals with “other” shift work (split/irregular/on-call/rotating); and lower in individuals who perceived their village wealth or spirit to be poor/average or weak/average, respectively. With the exception of potential sex-specific differences in WHIIRS scores based on diabetes status, perceived social, and sleep duration, the results were generally concordant between sexes (Table S4). The correlation between ESS and WHIIRS scores was Pearson R=−0.08 (*p*=0.08, Figure S1).

### 3.2 Variable selection and sleep outcome modeling

#### 3.2.1 ESS

In modeling the ESS, all variables listed in Table 2 were considered in the variable selection algorithm. In addition to the variables forced into all models (age, sex, BMI, rs373863828 genotype, and census region), the backward elimination approach selected a model including self-reported physical activity, work schedule, material lifestyle score, perceived stress, asthma, self-reported physical health, abdominal circumference, chronotype, children at home, and perceived village spirit as independent variables (Table 2). Of the variables selected, census region, physical activity, work schedule, material lifestyle score, perceived stress, and asthma diagnosis were significantly associated with the ESS. Specifically, ESS scores were higher in individuals who reported >0 minutes of MVPA than those reporting 0 minutes of MVPA (Est=1.88; 95% CI=1.12 to 2.75), individuals with higher material lifestyle scores (Est=0.18 per unit change; 95% CI=0.09 to 0.28), and individuals with asthma compared to those without (Est=2.85; 95% CI=1.25 to 4.51). ESS scores were lower in individuals residing in NWU compared with AUA (Est=−1.43; 95% CI=−2.39 to −0.41), individuals who reported no work compared with day shift work (Est=−2.26; 95% CI=−3.07 to −1.41), and individuals who reported higher perceived stress (Est=−0.14; 95% CI=−0.23 to −0.06] per unit change). RMSD ratios >1 underscored inflation for some variables with lower selection certainty, though this is expected in complex human data sets [31]. The relative conditional bias was negligible for variables with an inclusion frequency >90%, but more apparent in variables with lower selection certainty. Of note, the final bootstrap estimates resembled the selected model and global model estimates for all but the sex variable, indicating very little selection bias in the final aggregated model.

**Table 2.** Epworth Sleepiness Scale (ESS, possible scores of 0 to 24): Global model, selected model, and bootstrap-derived quantities for multi-model inference and assessment of model uncertainty

| Predictors | Global model |  |  | Bootstrap inclusion frequency (%) | Selected model |  |  | RMSD ratio | Relative conditional bias (%) | Final bootstrap estimates |  |  |
| --- | --- | --- | --- | --- | --- | --- | --- | --- | --- | --- | --- | --- |
|  | Est | 2.5th % | 97.5th % |  | Est | 2.5th % | 97.5th % |  |  | Est | 2.5th % | 97.5th % |
| (Intercept) | 3.19 | -3.87 | 10.26 |  | 6.89 | 1.76 | 12.02 | 1.14 | 28.26 | 4.06 | -3.41 | 12.47 |
| Age (years) | 0.03 | -0.01 | 0.07 | 100 (forced) | 0.02 | -0.02 | 0.05 | 1.06 | -9.10 | 0.03 | -0.01 | 0.07 |
| Sex, Female (Ref=Male) | 0.17 | -0.68 | 1.01 | 100 (forced) | 0.05 | -0.65 | 0.76 | 0.99 | -8.07 | 0.15 | -0.67 | 0.99 |
| Body mass index (kg/m<sup>2</sup>) | -0.16 | -0.28 | -0.03 | 100 (forced) | -0.16 | -0.28 | -0.04 | 1.18 | -6.85 | -0.15 | -0.27 | 0.03 |
| rs373863828 genotype (minor allele=A) | 0.34 | -0.13 | 0.81 | 100 (forced) | 0.39 | -0.06 | 0.84 | 1.02 | 8.89 | 0.37 | -0.09 | 0.84 |
| **Census Region, NWU/periurban (Ref=AUA/urban)** | **-1.41** | **-2.32** | **-0.51** | **100 (forced)** | **-1.50** | **-2.38** | **-0.61** | **1.05** | **0.51** | **-1.43** | **-2.39** | **-0.41** |
| Census Region, ROU/rural (Ref=AUA/urban) | -0.24 | -1.18 | 0.71 | 100 (forced) | -0.34 | -1.25 | 0.57 | 1.11 | 9.21 | -0.28 | -1.25 | 0.77 |
| **MVPA minutes daily, >0 minutes (Ref=0 minutes)** | **1.88** | **1.06** | **2.70** | **100** | **1.93** | **1.13** | **2.74** | **0.98** | **1.05** | **1.88** | **1.12** | **2.75** |
| Work Schedule, Afternoon/night shift (Ref=Day shift) | 1.13 | -0.63 | 2.88 | 100 | 1.15 | -0.53 | 2.84 | 0.98 | 5.08 | 1.18 | -0.52 | 2.84 |
| Work Schedule, Other [split/irregular] (Ref=Day shift) | -0.03 | -1.22 | 1.17 | 100 | 0.15 | -0.95 | 1.25 | 1.03 | -1.66 | 0.06 | -1.25 | 1.25 |
| **Work Schedule, Does Not Work (Ref=Day shift)** | **-2.28** | **-3.05** | **-1.51** | **100** | **-2.29** | **-3.02** | **-1.57** | **1.09** | **-1.11** | **-2.26** | **-3.07** | **-1.41** |
| **Material lifestyle score (total number of household assets)** | **0.18** | **0.09** | **0.27** | **99.6** | **0.19** | **0.10** | **0.27** | **1.05** | **1.96** | **0.18** | **0.09** | **0.28** |
| **Cohen's Perceived Stress Scale** | **-0.15** | **-0.23** | **-0.07** | **99.1** | **-0.15** | **-0.22** | **-0.08** | **1.12** | **-2.20** | **-0.14** | **-0.23** | **-0.06** |
| **Asthma, Yes (Ref=No)** | **2.92** | **1.27** | **4.56** | **98.2** | **2.76** | **1.16** | **4.35** | **1.02** | **0.38** | **2.85** | **1.25** | **4.51** |
| SF-8 Physical Health Component Score | -0.07 | -0.11 | -0.03 | 95.9 | -0.07 | -0.11 | -0.03 | 1.16 | 3.35 | -0.07 | -0.11 | 0 |
| Abdominal circumference (cm) | 0.09 | 0.03 | 0.14 | 90.6 | 0.08 | 0.03 | 0.14 | 1.27 | 0.88 | 0.08 | 0 | 0.14 |
| Chronotype, Evening person (Ref=Morning person) | 0.81 | 0.12 | 1.49 | 85.1 | 0.86 | 0.19 | 1.53 | 1.20 | 15.08 | 0.84 | 0 | 1.54 |
| Children at home, Yes (Ref=No) | 1.24 | 0.18 | 2.29 | 83.8 | 1.10 | 0.07 | 2.12 | 1.18 | 11.93 | 1.24 | 0 | 2.19 |
| Perceived village spirit, Weak or Average (Ref=Strong) | 0.70 | -0.12 | 1.52 | 61.8 | 0.66 | -0.10 | 1.43 | 1.23 | 31.97 | 0.67 | 0 | 1.45 |
| Multidimensional Scale of Perceived Social Support | 0.52 | -0.17 | 1.22 | 57.1 |  |  |  | 1.21 | 46.10 | 0.53 | 0 | 1.22 |
| SF-8 Mental Health Component Score | -0.03 | -0.07 | 0.01 | 54.9 |  |  |  | 1.25 | 71.01 | -0.03 | -0.08 | 0 |
| Uses sleeping pills, Yes (Ref=No) | -0.96 | -2.55 | 0.62 | 44.6 |  |  |  | 1.25 | 71.68 | 0 | -2.76 | 0 |
| Women's Health Initiative Insomnia Rating Scale | -0.08 | -0.21 | 0.05 | 43.6 |  |  |  | 1.18 | 70.12 | 0 | -0.22 | 0 |
| Hypertension, Yes (Ref=No) | 0.32 | -0.41 | 1.05 | 33.9 |  |  |  | 1.07 | 122.58 | 0 | 0 | 1.09 |
| Diabetes, Pre-diabetes (Ref=No) | -0.23 | -1.15 | 0.69 | 32.8 |  |  |  | 0.81 | 75.15 | 0 | -1.11 | 0.44 |
| Diabetes, Yes (Ref=No) | -0.57 | -1.63 | 0.48 | 32.8 |  |  |  | 1.10 | 81.18 | 0 | -1.56 | 0 |
| Perceived Social Conflict Score | 0.02 | -0.03 | 0.08 | 31.5 |  |  |  | 1.05 | 89.26 | 0 | -0.04 | 0.08 |
| Education (years) | 0.03 | -0.10 | 0.17 | 25.7 |  |  |  | 0.95 | 179.84 | 0 | -0.11 | 0.18 |
| Smokes, Yes (Ref=No) | 0.21 | -0.51 | 0.92 | 25.0 |  |  |  | 0.93 | 147.66 | 0 | -0.53 | 0.92 |
| Consumes alcohol, Yes (Ref=No) | 0.16 | -0.77 | 1.09 | 22.9 |  |  |  | 0.86 | 261.13 | 0 | -0.70 | 1.12 |
| Relationship status, Partnered (Ref=Not partnered) | -0.14 | -1.02 | 0.75 | 22.0 |  |  |  | 0.88 | 155.47 | 0 | -1.02 | 0.85 |
| Sleep duration (self-reported hours/weeknight or weekday) | 0.02 | -0.18 | 0.21 | 20.0 |  |  |  | 0.86 | -94.56 | 0 | -0.22 | 0.21 |
| Owns or rents home, Rents (Ref=Owns) | -0.05 | -1.59 | 1.50 | 19.9 |  |  |  | 0.84 | -616.88 | 0 | -1.60 | 1.71 |

Total number of participants, n=443; global and selected models presented alongside the final bootstrap model to assess the stability of the estimates and variable selection bias; global model shows the estimates and standard errors when all variables of interest are included in the model; selected model, selected via backward elimination with a significance level of 0.157 (AIC selection); final bootstrap estimates, indicate the bootstrap median estimate and the 2.5th and 97.5th percentiles interpreted as limits of 95% confidence intervals obtained by resampling-based multi-model inference; forced, indicates variables included in all models based on *a priori* knowledge of the Samoan population or to adjust for study design strategy; estimated shrinkage factor of model 0.905; selected model frequency 0.2%; all variance inflation factors of selected model <1.3, with the exception of BMI and abdominal circumference, with variance inflation factors of 2.7 and 2.6, respectively; Est, estimate; 2.5th and 97.5th percentiles interpreted as limits of 95% confidence intervals; RMSD, root mean squared difference; global model, R<sup>2</sup>=36.0, adjusted R<sup>2</sup>=30.9; selected model, R<sup>2</sup>=34.7, adjusted R<sup>2</sup>=31.8; bolded values indicate statistical significance based on bootstrap confidence intervals

#### 3.2.2 WHIIRS

In modeling the WHIIRS all variables listed in Table 3 were considered in the variable selection algorithm. In addition to the forced variables (described above), the backward elimination approach selected work schedule, perceived village wealth, perceived social support, children at home, asthma diagnosis, home ownership status, perceived village spirit, and use of sleeping pills as independent variables (Table 3). Of the selected variables, only work schedule and perceived village wealth were significantly associated with the WHIIRS. Specifically, WHIIRS scores were lower in individuals who reported “other” work schedules (i.e., split, irregular/on-call, or rotating shifts) compared with day shift work (Est=−1.96; 95% CI=−2.89 to −1.14) and individuals who perceived their village’s wealth to be poor/average compared with wealthy (Est=−0.94; 95% CI=−1.50 to −0.34). Model diagnostics were similar to those described above, with the exception of generally higher relative conditional bias.

**Table 3.** Women’s Health Initiative Insomnia Rating Scale (WHIIRS, possible scores 0 to 20): Global model, selected model, and bootstrap-derived quantities for multi-model inference and assessment of model uncertainty

| Predictors | Global model |  |  | Bootstrap inclusion frequency (%) | Selected model |  |  | RMSD ratio | Relative conditional bias (%) | Final bootstrap estimates |  |  |
| --- | --- | --- | --- | --- | --- | --- | --- | --- | --- | --- | --- | --- |
|  | Est | 2.5th % | 97.5th % |  | Est | 2.5th % | 97.5th % |  |  | Est | 2.5th % | 97.5th % |
| (Intercept) | 1.58 | -3.68 | 6.85 |  | 2.86 | -0.27 | 5.98 | 1.01 | 34.32 | 2.14 | -3.07 | 7.17 |
| Age (years) | -0.002 | -0.03 | 0.03 | 100 (forced) | 0.00 | -0.03 | 0.02 | 1.11 | 38.64 | 0.00 | -0.04 | 0.03 |
| Sex, Female (Ref=Male) | -0.06 | -0.69 | 0.58 | 100 (forced) | 0.11 | -0.38 | 0.61 | 1.01 | -38.71 | -0.03 | -0.70 | 0.56 |
| Body mass index (kg/m<sup>2</sup>) | 0.04 | -0.06 | 0.13 | 100 (forced) | 0.06 | 0.02 | 0.10 | 0.92 | 6.17 | 0.05 | -0.07 | 0.11 |
| rs373863828 genotype (minor allele=A) | -0.07 | -0.43 | 0.28 | 100 (forced) | -0.10 | -0.43 | 0.24 | 1.06 | 28.36 | -0.09 | -0.47 | 0.27 |
| Census Region, NWU (Ref=AUA) | 0.50 | -0.19 | 1.18 | 100 (forced) | 0.52 | -0.13 | 1.17 | 1.05 | 0.55 | 0.51 | -0.22 | 1.21 |
| Census Region, ROU (Ref=AUA) | -0.49 | -1.20 | 0.21 | 100 (forced) | -0.48 | -1.16 | 0.19 | 0.97 | 0.42 | -0.49 | -1.17 | 0.15 |
| Work Schedule, Afternoon/night shift (Ref=Day shift) | -0.05 | -1.36 | 1.26 | 99.5 | -0.25 | -1.49 | 1.00 | 1.07 | 170.07 | -0.13 | -1.49 | 1.23 |
| **Work Schedule, Other (Ref=Day shift)** | **-1.99** | **-2.86** | **-1.12** | **99.5** | **-1.74** | **-2.56** | **-0.93** | **1.06** | **-0.95** | **-1.96** | **-2.89** | **-1.14** |
| Work Schedule, Does Not Work (Ref=Day shift) | -0.39 | -0.99 | 0.20 | 99.5 | -0.13 | -0.66 | 0.39 | 1.10 | -18.58 | -0.30 | -0.95 | 0.29 |
| **Perceived village wealth, Poor or Average (Ref=Wealthy)** | **-0.89** | **-1.43** | **-0.35** | **97.5** | **-0.92** | **-1.44** | **-0.40** | **1.13** | **7.16** | **-0.94** | **-1.50** | **-0.34** |
| Multidimensional Scale of Perceived Social Support | 0.53 | 0.02 | 1.05 | 72.3 | 0.48 | -0.01 | 0.96 | 1.28 | 19.16 | 0.51 | 0 | 1.04 |
| Children at home, Yes (Ref=No) | 0.78 | -0.002 | 1.57 | 69.2 | 0.63 | -0.13 | 1.38 | 1.31 | 22.56 | 0.74 | 0 | 1.59 |
| Asthma, Yes (Ref=No) | 1.13 | -0.11 | 2.37 | 64.0 | 1.18 | 0.00 | 2.37 | 1.42 | 34.94 | 1.08 | 0 | 2.59 |
| Owens or rents home, Rents (Ref=Owns) | -0.95 | -2.10 | 0.20 | 61.5 | -0.90 | -2.02 | 0.22 | 1.28 | 38.68 | -0.94 | -2.18 | 0 |
| Perceived village spirit, Weak or Average (Ref=Strong) | -0.43 | -1.04 | 0.18 | 55.6 | -0.55 | -1.12 | 0.02 | 1.23 | 58.34 | -0.47 | -1.10 | 0 |
| Uses sleeping pills, Yes (Ref=No) | -0.97 | -2.15 | 0.21 | 55.5 | -0.87 | -2.01 | 0.27 | 1.25 | 33.86 | -0.87 | -2.09 | 0 |
| Perceived Social Conflict Score | 0.03 | -0.01 | 0.08 | 55.4 |  |  |  | 1.27 | 45.20 | 0.03 | 0 | 0.08 |
| Diabetes, Pre-diabetes (Ref=No) | 0.47 | -0.21 | 1.15 | 49.4 |  |  |  | 1.25 | 45.13 | 0 | 0 | 1.18 |
| Diabetes, Yes (Ref=No) | 0.61 | -0.18 | 1.39 | 49.4 |  |  |  | 1.35 | 53.11 | 0 | 0 | 1.48 |
| Relationship status, Partnered (Ref=Not partnered) | -0.43 | -1.09 | 0.22 | 45.9 |  |  |  | 1.27 | 65.15 | 0 | -1.15 | 0 |
| Consumes alcohol, Yes (Ref=No) | -0.42 | -1.11 | 0.27 | 42.7 |  |  |  | 1.13 | 68.99 | 0 | -1.06 | 0 |
| Epworth sleepiness scale | -0.04 | -0.12 | 0.03 | 41.8 |  |  |  | 1.15 | 59.96 | 0 | -0.11 | 0 |
| SF-8 Mental Health Component Score | 0.01 | -0.02 | 0.04 | 35.9 |  |  |  | 1.05 | 123.05 | 0 | 0 | 0.05 |
| Material lifestyle score (total number of household assets) | 0.03 | -0.04 | 0.10 | 35.1 |  |  |  | 1.04 | 115.97 | 0 | 0 | 0.10 |
| Hypertension, Yes (Ref=No) | -0.20 | -0.75 | 0.34 | 32.6 |  |  |  | 0.99 | 136.99 | 0 | -0.75 | 0 |
| Cohen's Perceived Stress Scale | -0.02 | -0.07 | 0.04 | 29.8 |  |  |  | 0.97 | 158.80 | 0 | -0.08 | 0.05 |
| Smokes, Yes (Ref=No) | 0.14 | -0.40 | 0.67 | 26.2 |  |  |  | 0.94 | 150.35 | 0 | -0.45 | 0.67 |
| Abdominal circumference (cm) | 0.01 | -0.03 | 0.05 | 22.4 |  |  |  | 0.88 | 306.37 | 0 | -0.03 | 0.05 |
| SF-8 Physical Health Component Score | -0.001 | -0.03 | 0.03 | 22.2 |  |  |  | 0.87 | -711.65 | 0 | -0.03 | 0.03 |
| Education (years) | 0.02 | -0.08 | 0.12 | 21.5 |  |  |  | 0.85 | 199.77 | 0 | -0.08 | 0.12 |
| MVPA minutes daily, >0 minutes (Ref=0 minutes) | 0.08 | -0.55 | 0.70 | 21.0 |  |  |  | 0.86 | 161.74 | 0 | -0.57 | 0.72 |
| Sleep duration (self-reported hours/weeknight or weekday) | -0.001 | -0.15 | 0.14 | 20.1 |  |  |  | 0.85 | 1746.24 | 0 | -0.15 | 0.15 |

Total number of participants, n=443; global and selected models presented alongside the final bootstrap model to assess the stability of the estimates and variable selection bias; global model shows the estimates and standard errors when all variables of interest are included in the model; selected model, selected via backward elimination with a significance level of 0.157 (AIC selection); final bootstrap estimates, indicate the bootstrap median estimate and the 2.5th and 97.5th percentiles interpreted as limits of 95% confidence intervals obtained by resampling-based multi-model inference; forced, indicates variables included in all models based on *a priori* knowledge of the Samoan population or to adjust for study design strategy; estimated shrinkage factor of model 0.796; selected model frequency 0.1%; all variance inflation factors of selected model <1.1; Est, estimate; 2.5th and 97.5th percentiles interpreted as limits of 95% confidence intervals; RMSD, root mean squared difference; global model, R<sup>2</sup>=19.9, adjusted R<sup>2</sup>=13.4; selected model, R<sup>2</sup>=17.6, adjusted R<sup>2</sup>=14.5; bolded values indicate statistical significance based on bootstrap confidence intervals

## 4. DISCUSSION

The prevalence of daytime sleepiness and insomnia – as well as factors associated with these measures – differed in this study compared with high-income countries, highlighting a critical gap in knowledge about sleep health in low- and middle-income settings. Specifically, with the exception of Maori individuals from New Zealand [32, 33], Samoan adults in this study had a generally higher prevalence, 20.0%, of excessive daytime sleepiness (ESS>10) compared with individuals from high-income countries. Comparatively, in a sample of older US adults, excessive daytime sleepiness prevalence by self-identified race/ethnicity was 18.8% in Black individuals, 13.6% in Hispanic individuals, 11.4% in White individuals, and 11.1% in Chinese individuals [4]. In a large community-based study of healthy individuals in New Zealand, the majority of whom were of European ancestry (81%), only 8.8% of individuals were found to have excessive daytime sleepiness [34], though this report is not population-representative as it is from blood donors (i.e., healthy donor effect). In a stratified random sample of Maori and non-Maori adults in New Zealand, however, 14.8% of participants reported an ESS>10 with excessive daytime sleepiness significantly higher in Maori individuals (21.3%) than non-Maori (13.9%) [33], a prevalence similar to that observed in the current study. A similar finding was observed in a larger stratified random sample of adults in New Zealand, in which ESS>10 were observed in 23.7% of Maori individuals compared with only 13.9% of non-Maori individuals [32]. In contrast, study participants had a lower prevalence of insomnia, 6.3%, compared with individuals from high-income countries. For example, in the same US study described above, insomnia (WHIIRS>10) prevalence was 26.5% in Hispanic individuals, 25.0% in Black individuals, 22.5% in White individuals, and 16.0% in Chinese individuals [4]. Of healthy individuals in the New Zealand study, 20.4% met criteria for primary insomnia based on the Diagnostic and Statistical Manual of Mental Disorders (DSM-IV) and International Classification of Sleep Disorders [34].

Of note, work schedule was the only variable significantly associated with both the ESS and WHIIRS. In multivariable analyses, day shift workers reported higher daytime sleepiness than those who did not work, as well as greater insomnia compared with those who worked “other” shifts (i.e., split, irregular/on-call, rotating). This latter observation contrasts with observations in high-income countries in which insomnia is more frequently observed in shift workers [19]. In *post hoc* analyses comparing responses to each of the four questions that make up the WHIIRS between day shift and other shift workers, the observation appears to be driven by problems of sleep maintenance rather than sleep latency, either reflecting impact of homeostatic drive or an environmental disturbance in day shift workers (e.g., schedule clashes between household members).

In the ESS model, along with work schedule, asthma diagnosis and self-reported physical activity had the strongest associations with daytime sleepiness. A relationship between sleepiness and asthma has been reported previously, and is attributed to potential sleep disruption from nocturnal bronchospasm or gastro-esophageal reflux, or to co-morbidities such as obesity and sleep apnea [35]. Very little data has been collected on asthma prevalence in Samoa and, like many low-and middle-income settings, it is likely underdiagnosed. It would be worth investigating whether asthma plays a role in other sleep disturbances in Samoa. Unexpectedly, sleepiness was also more common in individuals reporting higher levels of daily physical activity, which counters the notion that physical activity improves sleep health and reduces symptoms of daytime sleepiness [36]. In contrast to high-income countries where physical activity is often a leisure activity, physical activity in Samoa is traditionally work-related. Differences in the impact of leisure and work-related physical activity on sleep have been reported in Hispanic/Latino adults in the US, with shorter sleep duration reported in relationship to work-related but not leisure-related physical activity [37]. Perceived physical activity in Samoa tends to be highest among those engaged in subsistence farming and fishing activities, which often begin early in the morning to avoid the heat of the later day [38]. This may explain our observation – particularly if daily wake time and chronotype are mismatched. These findings should, however, be interpreted with caution given the usually imperfect correlation between perceived and observed physical activity in this setting [39].

Other variables associated with ESS scores included census region, perceived stress, and material lifestyle scores. In the absence of collinearity with social variables or physical activity, additional research is needed to better understand whether the lower daytime sleepiness observed in the periurban NWU region compared to the urban AUA region is due to differences in rest-activity-rhythms, occupation/commuting, differences in ambient lighting, and/or additional unobserved factors between geographic regions. The observation of less sleepiness in individuals reporting greater perceived stress conflicts with existing literature in high-income countries suggesting a positive association between stress and sleepiness [40]. A potential explanation could be that higher stress may result in hypervigilance, which in these communities may have alerting effects that counter the negative effects of poor sleep quality typically associated with stress. This hypervigilance theory would likely result in more insomnia symptoms, however, which was not observed. The observation of higher daytime sleepiness in individuals with higher material lifestyle scores (suggesting greater socioeconomic resources) is also in direct contrast with research in New Zealand in which greater daytime sleepiness was observed in lower income individuals [32]. It is possible that this observation is due to differences in societal support between the two countries. For example, in Samoa, support networks are very strong in communities with traditional culture, which could decrease the amount of stress traditionally associated with fewer economic resources in high-income countries. Alternatively, there could be greater expectations from the community on those with, or having the appearance of, greater socioeconomic resources (e.g., *fa’alavelave*, offering financial support to a family, village, or church event such as a funeral), leading to higher stress and poorer sleep health.

In the WHIIRS-specific model, the only variables that had a statistically significant association with insomnia included work schedule as discussed above, and perceived village wealth. Specifically, lower WHIIRS scores were observed in individuals who perceived their village wealth to be poor/average. This observation, paired with perceived village spirit (which had the same direction of association but was not statistically significant), generally conflicts with research in high-income countries. For example, in the US, adverse neighborhood factors have been associated with a higher prevalence of insomnia in a Hispanic/Latino population [41] and low neighborhood social cohesion has been associated with shorter sleep duration in Native Hawaiian and Pacific Islanders [6]. Qualitative research is needed to better understand the perceived village wealth and spirit variables examined here, including their potential utility for improving health and sleep outcomes.

Other unexpected findings surrounded adiposity measures, sleep duration, and rs373863828 genotype. While adiposity measures were associated with WHIIRS scores in bivariate analyses, these associations did not persist in multivariable modeling and were not observed for ESS across the study. It is possible that because the mean BMI and abdominal circumference of the sample was so high, there was some degree of obesity-related sleep disturbance for all participants. Likewise, in the multivariable analyses, sleep duration was not found to be important in explaining sleep outcomes in this sample which was unexpected. Even when evaluating the typical ‘u-shaped’ distribution of sleep duration by categorizing individuals reporting short sleep (<6 hours), sufficient sleep (6-9 hours), or long sleep (>9 hours) (as well as a variety of other cutoff points in *post hoc* analyses), there was no difference in the ESS scores between groups in bivariate analyses (Table S3). In contrast, we did observe marginally lower WHIIRS scores in the short (< 6 hours) and long sleep (>9 hours) groups in men, but not women (Table S4). Finally, *CREBRF* rs373863828 genotype was found to be significantly associated with ESS scores in bivariate but not multivariable analyses controlling for BMI, abdominal circumference, and diabetes status. While regression diagnostics suggest no collinearity between genotype and adiposity measures, it is possible that the inclusion of these other variables absorbs variation related to genotype, but no substantive conclusion can be made.

Of note, strengths of this study included the focus on Samoan adults, an underrepresented population in sleep research specifically and health research more generally, and a rigorous variable selection and analytical approach, providing de-biased multi-model inferences. Weaknesses included the cross-sectional and descriptive nature of this study and the non-representative sampling approach taken for the 2017-2019 wave of data collection described above, which potentially limits the generalizability of the presented findings.

## 5. CONCLUSION

Adults in Samoa have a generally higher prevalence of excessive daytime sleepiness, but lower prevalence of insomnia, compared with individuals from high-income countries. Participant factors, particularly work schedule, self-reported physical activity, asthma, and geographic region of residence showed strong associations with daytime sleepiness and may assist in identifying those at risk for daytime sleepiness. Many observations differed from those in high-income countries underscoring vast potential cultural or environmental differences in the drivers of sleep health. Given the role of sleep in metabolic health, and the unexpected differences observed here, more research and contextually targeted interventions to improve sleep practices in this already high-risk population are needed.

## Data Availability

To be deposited in dbGAP under accession #phs000914.v1.p1.

https://www.ncbi.nlm.nih.gov/projects/gap/cgi-bin/study.cgi?study_id=phs000914.v1.p1

## Abbreviations

CREBRF: CREB3 Regulatory Factor
ESS: Epworth Sleepiness Scale
WHIIRS: Women’s Health Initiative Insomnia Rating Scale
AUA: Apia urban area (urban)
NWU: Northwest ‘Upolu (periurban)
ROU: Rest of ‘Upolu (rural)
BMI: Body mass index
HbA1c: Hemoglobin A1c
BP: Blood pressure
MVPA: moderate to vigorous physical activity
SF-8: Short form 8
MSPSS: Multidimensional scale of perceived social support
AIC: Akaike information criterion
EPV_global_: Events per variable
ANOVA: One-way analysis of variance
CI: Confidence intervals
RMSD: Root mean squared difference
SI: Supplementary information

## DECLARATIONS

## Acknowledgements

We would like to thank the participants for their involvement in this research as well as the local village authorities, the Samoa Ministry of Health, the Samoa Bureau of Statistics, and the Ministry of Women, Community and Social Development for their support of this work. A particular *fa’afetai tele lava* to our research assistants – Melania Selu, Vaimoana Lupematisilia, Folla Unasa, and Lupesina Vesi – without whom this work would not be possible.

## Ethical Approval

This study was approved by the Institutional Review Boards (IRB) at Yale and Brown Universities, with Yale University serving as the IRB of record (#1604017547). Data analysis activities at the University of Pittsburgh were reviewed by their IRB and were determined to be exempt (IRB #PRO16040077) based on their receipt of only deidentified data. Activities of the Sleep Reading Center were approved by Mass General Brigham (formerly Partners Healthcare). The study was also approved by the Health Research Committee of the Samoa Ministry of Health.

## Consent to Participate

Written informed consent was obtained from all participants prior to enrollment.

## Availability of Data

To be deposited in dbGAP under accession #phs000914.v1.p1.

## Authors’ Roles

LWH, JCC, STM, BEC, SR, and NLH conceived and designed the present study with additional methodological feedback from all authors. LWH performed all statistical analyses with oversight and methodological guidance from JCC and DEW. LWH wrote the first draft of the manuscript and critically revised based on co-author feedback. NLH supervised all aspects of the project. AP was responsible for overseeing data collection activities with supervision from NLH, STM, TN, and MSR. SR and BC assisted NLH with designing sleep study questionnaires and provided oversight of sleep data acquisition. STM is the principal investigator of the parent study and, with NLH and DEW, was responsible for acquisition of the study funding. All authors contributed to the interpretation of the data and results. All authors reviewed, critically revised, and approved the final manuscript. All authors agree to be accountable for all aspects of the work.

