## Additional File for "Correlates of Daytime Sleepiness and Insomnia among Adults in Samoa"

***Supplementary Material, Additional File 1***

Lacey W. Heinsberg, PhD, RN*; Jenna C. Carlson, PhD;
Alysa Pomer, PhD; Brian E. Cade, PhD; Take Naseri, MBBS, MPH;
Muagututia Sefuiva Reupena, MS; Daniel E. Weeks, PhD; Stephen McGarvey, PhD;
Susan Redline, MD; Nicola L. Hawley, PhD

*Corresponding author
Department of Human Genetics
Graduate School of Public Health
University of Pittsburgh


**TABLE OF CONTENTS**

| Section | Table/Figure | Title | Page |
|---|---|---|---|
| *Section I* | | *Translated Abstract (Samoan)* | 2 |
| *Section II* | | *Expanded methods* | 3 |
| | Table S1 | Detailed variable definitions | 3 |
| | | Expanded statistical analyses | 6 |
| *Section III* | | *Expanded results* | 7 |
| | Table S2 | Imputation summary | 7 |
| | Table S3 | Bivariate associations between participant characteristics and the Epworth Sleepiness Scale (ESS) | 8 |
| | Table S4 | Bivariate associations between participant characteristics and the Women's Health Initiative Insomnia Rating Scale (WHIIRS) | 10 |
| | Figure S1 | Scatterplot depicting correlation between the Epworth Sleepiness Scale (ESS) and the Women's the Health Initiative Insomnia Rating Scale (WHIIRS) | 12 |

**Section I. Translated Abstract**

Abstract translated to Samoan available upon request.

**Section II. Expanded Methods**

**Table S1.** Detailed variable definitions

| *Biological factors and health history* | | |
|---|---|---|
| *Variable* | *Original questionnaire response or variable measurement* | *Summary of variable construction/recoding for the present study* |
| Age (years) | Date of birth used to compute age in years | No modification made, treated as a continuous variable |
| Body mass index (kg/m$^2$) | Weight and height were measured in duplicate to the nearest 0.1 kg and 0.1 cm, averaged, and used to compute body mass index in kg/m2 | No modification made, treated as a continuous variable |
| Abdominal circumference (cm) | Abdominal circumference was measured in duplicate to the nearest 0.1 cm at the level of the umbilicus and averaged | No modification made, treated as a continuous variable |
| Diabetes (no/pre-diabetes/yes) | Hemoglobin A1c (HbA1c) values measured from fasting blood samples<br>Diabetes medication self-reported by participants | Diabetes status was determined based on current use of diabetes medication or HbA1c values and defined as no diabetes (no use of diabetes medication and HbA1c <5.7%), pre-diabetes (no use of diabetes medication and HbA1c of 5.7-6.4%), or diabetes present (use of diabetes medication and/or HbA1c>6.4%) |
| Hypertension (yes/no) | Blood pressure was measured in triplicate and the second and third values were averaged for use in analyses<br>Hypertension medication self-reported by participants | Hypertension was defined as an average value >140/90 mmHg and/or current hypertension medication use |
| Asthma | A history of asthma diagnosis was self-reported by participants (yes/no) | No modification made, treated as categorical variable (yes/no) |
| SF-8 Physical Health Component Score (possible scores of 0-100) | The Short Form (SF) 8 was used to assess an individual's perceived physical health using the Physical Health Component Score which results in scores ranging from 0 to 100 with higher scores indicating better perceived health | No modification made, treated as a continuous variable |
| SF-8 Mental Health Component Score (possible scores of 0-100) | The Short Form (SF) 8 was used to assess an individual's perceived mental health using the Mental Health Component Score which results in scores ranging from 0 to 100 with higher scores indicating better perceived health | No modification made, treated as a continuous variable |
| *Sociodemographic and behavioral factors* | | |
| *Variable* | *Original questionnaire response or variable measurement* | *Summary of variable construction/recoding for the present study* |
| Census region (AUA/NWU/ROU) | In which census region does the participant reside? (AUA/NWU/ROU) | No modification made, treated as categorical variable (AUA/NWU/ROU) |
| Material lifestyle score (total number of household assets, possible scores of 0-18) | A material lifestyle score used as a proxy for socioeconomic status in Samoa was computed as the total number of self-reported items owned from a household inventory list (i.e., refrigerator, freezer, electricity in house, portable stereo/MP3 player, microwave oven, rice cooker, blender, sewing machine, television, VCR or DVD player, couch, washing machine, mobile telephone, computer or laptop, tablet computer, electric fan, air conditioner, and motorized vehicle). | No modification made, treated as a continuous variable |
| Education (years) | Self-reported by participants as the total years of completed education | No modification made, treated as a continuous variable |
| Relationship status (partnered/not partnered) | Self-reported response of:<br>Currently Married<br>Cohabitating<br>Never married<br>Separated<br>Divorced<br>Widowed | Recoded as:<br>Partnered (currently married or cohabitating)<br>Not partnered (never married, separated, divorced, or widowed) |
| Children at home (yes/no) | Total number of children who share the house (continuous number) | Recoded as do children live in the home:<br>Yes (>=1 child sharing the home) |

| | | |
|---|---|---|
| | | No (0 children sharing the home) |
| Physical activity (0 MPVA minutes/<0 MVPA minutes) | Average number of minutes of moderate-vigorous physical activity (MVPA) per day (continuous number) measured using the Global Physical Activity Questionnaire | Dichotomized as 0/>0 MVPA minutes based on an extreme floor effect seen in this population |
| Work schedule (day shift/afternoon or night shift/other [split, irregular/on-call, or rotating]/does not work) | Self-reported response of: | Due to cell counts (i.e., afternoon=9, night=13, split=40, irregular/on-call=3, rotating=8), variable recoded as:<br>Day shift (day shift)<br>Afternoon or night shift (afternoon shift or night shift)<br>Other (split, irregular/on-call, or rotating shifts)<br>Does not work (Don't work)<br><br>Note: Bivariate analyses showed no difference in ESS/WHIIRS within collapsed categories |
| | Day shift | |
| | Afternoon shift | |
| | Night shift | |
| | Split shift | |
| | Irregular shift/on-call | |
| | Rotating shifts | |
| | Don't work | |
| Home ownership status (own/rent) | Self-reported by participants (owns/rents home) | No modification made, treated as categorical variable (owns/rents home) |
| Smokes (yes/no) | Participant smokes cigarettes, cigars, or pipes (self-report, yes/no) | No modification made, treated as categorical variable (yes/no) |
| Consumes alcohol (yes/no) | Self-reported response of how often has the participant had at least one drink in the last 12 months: | Dichotomized as consumption of alcohol in the last 12 months:<br>No (never)<br>Yes (any other response) |
| | Never | |
| | Five or more days a week | |
| | 1 to 4 days per week | |
| | 1 to 3 days per month | |
| | Less than once a month | |
| Uses sleeping pills (yes/no) | Self-reported response of how often has the participant used sleeping pills to help them sleep (in the past 4 weeks): | Dichotomized as sleeping pills to help them sleep in the past 4 weeks:<br>No (No, not in the past 4 weeks)<br>Yes (any other response) |
| | No, not in the past 4 weeks | |
| | Yes, less than once a week | |
| | Yes, 1 or 2 times a week | |
| | Yes, 3 or 4 times a week | |
| | Yes, 5 or more times a week | |
| Perceived village wealth (poor or average compared with wealthy) | Self-reported response of how the participant perceives the wealth of the village in which they live: | Dichotomized as:<br>Poor/average (very poor, poor, or average)<br>Wealthy (wealthy or very wealthy) |
| | Very poor | |
| | Poor | |
| | Average | |
| | Wealthy | |
| | Very wealthy | |
| Perceived village spirit (weak or average compared with strong) | Self-reported response of how the participant perceives the community spirit of the village in which they live: | Dichotomized as:<br>Weak/average (very weak, weak, or average)<br>Strong (strong or very strong) |
| | Very weak | |
| | Weak | |
| | Average | |
| | Strong | |

| | | |
|---|---|---|
| | Very strong | |
| Cohen's Perceived Stress Scale (possible scores of 0-40) | Perceived stress was measured using Cohen's Perceived Stress Scale which consists of 10 questions and ranges from 0 to 40 with higher scores indicating higher perceived stress | No modification made, treated as a continuous variable |
| Multidimensional Scale of Perceived Social Support (possible scores of 0-5) | Social support was measured using the total score of the Multidimensional Scale of Perceived Social Support which includes 12 questions related to support from family, friends, and an individual's significant other, and ranges from 1 to 5 with higher scores indicating more perceived support | No modification made, treated as a continuous variable |
| Perceived Social Conflict Score (possible scores of 6-30) | Perceived social conflict was measured using the Perceived Social Conflict Score which was calculated from 6 questions and ranges from 6 to 30 with higher scores indicating higher perceived social conflict | No modification made, treated as a continuous variable |
| *Sleep measures* | | |
| *Variable* | *Original questionnaire response or variable measurement* | *Summary of variable construction/recoding for the present study* |
| Epworth Sleepiness Scale (ESS, possible scores of 0 to 24) | The ESS ranges from 0 to 24 with higher scores indicating greater daytime sleepiness; self-reported and calculated via questionnaire | No modification made, treated as a continuous variable |
| Excessive daytime sleepiness (yes/no) | ESS described above | Recoded as excessive daytime sleepiness:<br>Yes (ESS>10)<br>No (ESS<=10) |
| Women's Health Initiative Insomnia Rating Scale (WHIIRS, possible scores of 0 to 20) | The WHIIRS ranges from 0 to 20 with higher scores indicating greater severity of insomnia; self-reported and calculated via questionnaire | No modification made, treated as a continuous variable |
| Insomnia (yes/no) | WHIIRS described above | Recoded based on WHIIRS levels that are reported to be consistent with clinical thresholds for insomnia:<br>Yes (WHIIRS>10)<br>No (WHIIRS<=10) |
| Chronotype (morning/evening) | Self-reported by participants using the question "One hears about 'morning' and 'evening' type people. Which one of these types do you consider yourself to be?", modified from the Horne-Ostberg questionnaire.[1] Self-reported response of:<br>Definitely a "morning" type<br>Rather more a "morning" than an "evening" type<br>Rather more an "evening" than a "morning" type<br>Definitely an "evening" type<br>Neither a "morning" or an "evening" type | Dichotomized as:<br>Morning (definitely a "morning" type or rather more a "morning" than an "evening" type)<br>Evening (definitely an "evening" type or rather more an "evening" type than a "morning" type)<br>"Neither" responses set to missing due to small cell count (n=12) |
| Sleep duration (hours/weeknight or weekday) | Self-reported by participant; Participant's usual bedtime on weekdays or work or school days and participant's usual wake time on weekdays or work or school days | Bed and wake times used to calculate average nightly sleep duration on weekdays or work or school days (hours) |
| Sleep duration categorical | Self-reported by participant; Participant's usual bedtime on weekdays or work or school days and participant's usual wake time on weekdays or work or school days | Categorized as:<br><6 hours (short sleep)<br>6-9 hours (sufficient sleep)<br>>9 hours (long sleep) |

**Expanded statistical analyses**

All statistical analyses were performed in R version 3.6.0.[34] Standard descriptive statistics were computed based on variable type including means, standard deviations (SD), medians, and minimum/maximum values for continuous variables and frequency counts and percentages for categorical variables. Data were examined graphically and statistically to identify outliers and assess patterns of missing data. Within-domain multivariate imputation by chained equations was performed using the mice package[35] in R for continuous summary score variables with a missingness of 5-10% (i.e., SF-8 Physical and Mental Health, material lifestyle score, Cohen's Perceived Stress Scale, the Multidimensional Scale of Perceived Social Support, Perceived Social Conflict Score, ESS, and WHIIRS) as detailed in the SI (Table S2), and sensitivity analyses were performed to assess the influence of imputation.

Preliminary analyses were performed to evaluate bivariate associations between participant factors and sleep outcomes (ESS and WHIIRS), including both graphical (e.g., scatter or sina plots) and statistical examination of the data. Depending on the variable type and distribution, relationships were evaluated using Pearson or Spearman correlations, t-tests, or one-way analysis of variance (ANOVA). Levene's test for homogeneity of variance was used, and in cases in which the assumption of constant variance was violated, Welch's one-way analysis of means was used to evaluate relationships between groups. Given sex-specific health differences in the Samoan population[18,19,36], and the role of sex in sleep health[37,38], bivariate analyses were performed both for the overall sample and stratified by sex. Based on the results of bivariate analyses, sex stratification was not carried forward for the multivariable modeling in order to retain the largest sample size possible, but sex was included in regression models.

As the first study to examine sleep health in the Samoan population, and with a lack of strong theory to support *a priori* modeling, multiple linear regression and backwards elimination with Akaike information criterion (AIC) was used for variable selection[39], treating ESS or WHIIRS as the outcome variables. Age, sex, BMI, census region, and rs373863828 genotype (using additive coding based on the total number of minor allele (A) copies) were included in all models (i.e., forced) based on *a priori* knowledge of the Samoan population or to adjust for study design. Additional variables of interest included those described above. Only individuals with complete data post-imputation for all 28 variables of interest could be included in the multivariable modeling (n=443), making the global model number of events per variable ($EPV_{global}$) 443/28=15.8. Given the potential for variable-selection bias and issues associated with the use of the same data set for both variable selection and post-selection inference, a bootstrap stability investigation was performed to both assess the impact of variable selection and perform multi-model inference as recommended by Heinze, Wallisch, and Dunkler.[39] As part of this process, the bootstrap-inclusion frequency was calculated for each variable of interest and variation in regression coefficients around the full model estimates was assessed using bootstrapped resamples, repeating the backwards elimination selection procedure 1,000 times.

Coefficients and 95% confidence intervals (CI) were reported for the global model (i.e., including all input variables of interest) and the selected model (i.e., including variables chosen by backwards elimination). Final multi-model coefficients and 95% CI used for inference were reported as the median of the bootstrap distribution with multiple comparison interpretation built into the modeling approach. The root mean squared difference (RMSD) ratio, relative conditional bias, and model selection frequencies were calculated to further assess variable-selection. The RMSD ratio was computed as the variable-specific RMSD of the bootstrap estimates divided by the standard error in the global model (assumed unbiased), representing the variance inflation (values >1) or variance deflation (values <1) consequent to variable selection. The relative conditional bias was calculated as the differences of the mean of resampled regression coefficients and the global model regression coefficient, divided by the global model regression coefficient, representing the bias present if a variable was selected because its regression coefficient appeared extreme in a specific resample. Model selection frequencies indicate how likely a particular set of variables were to be selected together. Selected model assessment was performed using residual analysis, collinearity, and influence diagnostics. To stabilize regression parameters, variables were collapsed as described in the main text and further detailed in Table S1 below.

**Section III. Expanded results**

**Table S2.** Imputation summary

| | Raw data, no imputation | | | Data post-imputation | | |
|---|---|---|---|---|---|---|
| | *Overall (n=519)* | *Males (n=233)* | *Females (n=286)* | *Overall (n=519)* | *Males (n=233)* | *Females (n=286)* |
| SF-8 Physical Health Component Score | | | | | | |
| Mean (SD) | 43.4 (8.4) | 43.7 (8.4) | 43.2 (8.4) | 43.4 (8.4) | 43.7 (8.3) | 43.2 (8.5) |
| Median [Min, Max] | 42.9 [17.4, 63.1] | 42.9 [17.4, 63.1] | 42.9 [17.9, 62.4] | 42.9 [17.4, 63.1] | 42.9 [17.4, 63.1] | 42.6 [17.9, 62.4] |
| Missing | 40 (7.7%) | 12 (5.2%) | 28 (9.8%) | 28 (5.4%) | 7 (3.0%) | 21 (7.3%) |
| SF-8 Mental Health Component Score | | | | | | |
| Mean (SD) | 46.8 (9.9) | 47.2 (10.0) | 46.6 (9.8) | 46.7 (9.8) | 47.1 (9.9) | 46.5 (9.7) |
| Median [Min, Max] | 44.9 [15.2, 66.7] | 46.2 [19.9, 65.5] | 44.6 [15.2, 66.7] | 44.6 [15.2, 66.7] | 45.3 [19.9, 65.5] | 44.5 [15.2, 66.7] |
| Missing | 40 (7.7%) | 12 (5.2%) | 28 (9.8%) | 28 (5.4%) | 7 (3.0%) | 21 (7.3%) |
| Material lifestyle score | | | | | | |
| Mean (SD) | 8.0 (4.0) | 8.1 (4.1) | 8.0 (3.9) | 8.0 (4.0) | 8.1 (4.2) | 7.9 (3.9) |
| Median [Min, Max] | 8 [0, 18.0] | 7 [1.0, 17.0] | 8 [0, 18.0] | 7 [0, 18.0] | 7.0 [1.00, 17.0] | 8.0 [0, 18.0] |
| Missing | 36 (6.9%) | 16 (6.9%) | 20 (7.0%) | 3 (0.6%) | 1 (0.4%) | 2 (0.7%) |
| Cohen's Perceived Stress Scale | | | | | | |
| Mean (SD) | 17.8 (5.5) | 17.1 (6.0) | 18.5 (5.0) | 17.8 (5.5) | 17.1 (6.0) | 18.4 (4.9) |
| Median [Min, Max] | 20.0 [0, 32.0] | 19.0 [0, 32.0] | 20.0 [0, 28.0] | 20.0 [0, 32.0] | 19.0 [0, 32.0] | 20.0 [0, 28.0] |
| Missing | 39 (7.5%) | 12 (5.2%) | 27 (9.4%) | 28 (5.4%) | 7 (3.0%) | 21 (7.3%) |
| Multidimensional Scale of Perceived Social Support | | | | | | |
| Mean (SD) | 3.9 (0.5) | 3.9 (0.5) | 3.9 (0.5) | 3.9 (0.5) | 3.9 (0.5) | 3.9 (0.5) |
| Median [Min, Max] | 4.0 [1.6, 5.0] | 4.0 [1.6, 5.0] | 4.0 [1.7, 5.0] | 4.0 [1.6, 5.0] | 4.0 [1.6, 5.0] | 4.0 [1.7, 5.0] |
| Missing | 52 (10.0%) | 20 (8.6%) | 32 (11.2%) | 28 (5.4%) | 7 (3.0%) | 21 (7.3%) |
| Perceived Social Conflict Score | | | | | | |
| Mean (SD) | 17.2 (5.9) | 16.9 (6.0) | 17.6 (5.9) | 17.3 (5.9) | 17.0 (6.0) | 17.6 (5.8) |
| Median [Min, Max] | 16.0 [6.0, 30.0] | 16.0 [6.0, 30.0] | 17.0 [6.0, 30.0] | 16.0 [6.0, 30.0] | 16.0 [6.0, 30.0] | 17.0 [6.0, 30.0] |
| Missing | 39 (7.5%) | 10 (4.3%) | 29 (10.1%) | 28 (5.4%) | 7 (3.0%) | 21 (7.3%) |
| Epworth Sleepiness Scale | | | | | | |
| Mean (SD) | 6.8 (4.1) | 7.3 (4.1) | 6.4 (4.1) | 6.9 (4.1) | 7.4 (4.1) | 6.5 (4.1) |
| Median [Min, Max] | 7.0 [0, 16.0] | 8.0 [0, 15.0] | 7.0 [0, 16.0] | 7.0 [0, 16.0] | 8.0 [0, 15.0] | 7.0 [0, 16.0] |
| Missing | 48 (9.2%) | 14 (6.0%) | 34 (11.9%) | 29 (5.6%) | 7 (3%) | 22 (7.7%) |
| Women's Health Initiative Insomnia Rating Scale | | | | | | |
| Mean (SD) | 6.5 (2.7) | 6.3 (2.7) | 6.6 (2.8) | 6.5 (2.7) | 6.3 (2.7) | 6.6 (2.8) |
| Median [Min, Max] | 6.0 [0, 16.0] | 6.0 [0, 16.0] | 6.0 [0, 15.0] | 6.0 [0, 16.0] | 6.0 [0, 16.0] | 6.0 [0, 15.0] |
| Missing | 33 (6.4%) | 9 (3.9%) | 24 (8.4%) | 29 (5.6%) | 8 (3.4%) | 21 (7.3%) |

**Table S3.** Bivariate associations between participant characteristics and the Epworth Sleepiness Scale (ESS, possible scores of 0 to 24)

| *Biological factors and health history* | *Overall (n=519)* | | | *Males (n=233)* | | | *Females (n=286)* | | |
|---|---|---|---|---|---|---|---|---|---|
| | n | Cor Coef or Mean (SD) | *p* | n | Cor Coef or Mean (SD) | *p* | n | Cor Coef or Mean (SD) | *p* |
| Age (years) | 490 | -0.01 | 0.99[a] | 226 | 0.09 | 0.19[a] | 264 | -0.11 | 0.08[a] |
| Sex | 490 | | | NA | | | | | |
| Male | 226 | 7.4 (4.1) | **0.013**[b] | | | | | | |
| Female | 264 | 6.5 (4.1) | | | | | | | |
| Body mass index (kg/m$^2$) | 488 | 0.04 | 0.37[a] | 226 | 0.08 | 0.22[a] | 262 | 0.07 | 0.26[a] |
| Abdominal circumference (cm) | 488 | 0.08 | 0.07[a] | 226 | 0.12 | 0.07[a] | 262 | 0.09 | 0.14[a] |
| rs373863828 genotype | 490 | | | 226 | | | 264 | | |
| GG | 212 | 6.1 (4.3) | | 97 | 6.9 (4.2) | | 115 | 5.5 (4.2) | |
| AG | 192 | 7.1 (3.9) | **0.0002**[c] | 88 | 7.2 (3.9) | **0.025**[c] | 104 | 7.0 (3.9) | **0.004**[c] |
| AA | 86 | 8.2 (3.7) | | 41 | 8.9 (3.8) | | 45 | 7.5 (3.5) | |
| Diabetes | 490 | | | 226 | | | 264 | | |
| No | 91 | 7.1 (3.7) | | 49 | 7.9 (3.5) | | 42 | 6.1 (3.8) | |
| Pre-diabetes | 245 | 6.9 (4.0) | 0.73[d] | 111 | 7.2 (4.1) | 0.59[d] | 134 | 6.7 (3.9) | 0.64[d] |
| Yes | 154 | 6.7 (4.4) | | 66 | 7.2 (4.4) | | 88 | 6.3 (4.4) | |
| Hypertension | 480 | | | 225 | | | 255 | | |
| No | 322 | 6.8 (4.1) | 0.65[b] | 158 | 7.2 (4.1) | 0.23[b] | 164 | 6.5 (4.1) | 0.80[b] |
| Yes | 158 | 7.0 (4.1) | | 67 | 7.9 (4.0) | | 91 | 6.4 (4.0) | |
| Asthma | 488 | | | 224 | | | 264 | | |
| No | 465 | 6.8 (4.1) | **0.025**[b] | 211 | 7.3 (4.0) | 0.24[b] | 254 | 6.4 (4.1) | 0.06[b] |
| Yes | 23 | 8.8 (3.9) | | 13 | 8.9 (4.5) | | 10 | 8.7 (3.4) | |
| SF-8 Physical Health Component Score[f] | 490 | -0.11 | **0.012**[a] | 226 | -0.03 | 0.70[a] | 264 | -0.19 | **0.002**[a] |
| SF-8 Mental Health Component Score[f] | 490 | -0.08 | 0.07[a] | 226 | -0.04 | 0.56[a] | 264 | -0.13 | **0.037**[a] |
| *Sociodemographic and behavioral factors* | *Overall (n=519)* | | | *Males (n=233)* | | | *Females (n=286)* | | |
| | n | Cor Coef or Mean (SD) | *p* | n | Cor Coef or Mean (SD) | *p* | n | Cor Coef or Mean (SD) | *p* |
| Census region | 490 | | | 226 | | | 264 | | |
| AUA | 102 | 7.9 (4.1) | | 46 | 8.0 (4.2) | | 56 | 7.8 (4.1) | |
| NWU | 211 | 5.9 (3.7) | **6.32E-06**[d] | 101 | 6.5 (3.6) | **0.014**[d] | 110 | 5.3 (3.6) | **0.0001**[d] |
| ROU | 177 | 7.5 (4.3) | | 79 | 8.1 (4.3) | | 98 | 7.0 (4.2) | |
| Material lifestyle score (total number of household assets)[f] | 487 | 0.17 | **0.0001**[a] | 225 | 0.13 | **0.048**[a] | 262 | 0.21 | **0.0006**[a] |
| Education (years) | 489 | 0.04 | 0.39[a] | 225 | 0.06 | 0.39[a] | 264 | 0.05 | 0.42[a] |
| Relationship status | 489 | | | 226 | | | 263 | | |
| Partnered | 398 | 6.9 (4.1) | 0.68[b] | 186 | 7.3 (4.0) | 0.87[b] | 212 | 6.1 (3.4) | 0.46[b] |
| Not partnered | 91 | 6.7 (4.0) | | 40 | 7.5 (4.6) | | 51 | 6.5 (4.2) | |
| Children at home | 489 | | | 225 | | | 264 | | |
| No | 53 | 6.5 (4.1) | 0.48[b] | 25 | 8.5 (3.8) | 0.14[b] | 28 | 4.8 (3.6) | **0.014**[b] |
| Yes | 436 | 6.9 (4.1) | | 200 | 7.3 (4.1) | | 236 | 6.7 (4.1) | |
| Physical activity | 488 | | | 224 | | | 264 | | |
| 0 MVPA minutes/day | 359 | 6.2 (4.0) | **2.90E-11**[b] | 138 | 6.5 (4.1) | **0.0001**[b] | 221 | 5.9 (4.0) | **2.62E-07**[b] |
| >0 MVPA minutes/day | 129 | 8.8 (3.5) | | 86 | 8.6 (3.7) | | 43 | 9.1 (3.2) | |
| Work schedule | 489 | | | 225 | | | 264 | | |
| Day shift | 226 | 7.3 (3.4) | | 116 | 7.4 (3.7) | | 110 | 7.3 (3.1) | |
| Afternoon or night shift | 22 | 10.4 (2.8) | **7.75E-10**[d] | 11 | 10.9 (2.7) | **0.0002**[d] | 11 | 9.9 (3.0) | **6.43E-06**[d] |
| Other [split, irregular/on-call, rotating] | 51 | 8.3 (3.8) | | 18 | 9.3 (3.8) | | 33 | 7.8 (3.8) | |
| Does not work | 190 | 5.5 (4.5) | | 80 | 6.4 (4.4) | | 110 | 4.8 (4.5) | |
| Owns or rents home | 487 | | | 224 | | | 263 | | |
| Owns | 463 | 6.8 (4.1) | 0.38[b] | 217 | 7.4 (4.0) | 0.98[b] | 246 | 6.4 (4.1) | 0.13[b] |
| Rents | 24 | 7.5 (3.3) | | 7 | 7.4 (4.7) | | 17 | 7.5 (2.7) | |
| Smokes | 489 | | | 226 | | | 263 | | |
| No | 298 | 6.6 (4.1) | **0.038**[b] | 109 | 7.3 (4.2) | 0.79[b] | 189 | 6.2 (4.1) | 0.06[b] |
| Yes | 191 | 7.4 (4.0) | | 117 | 7.4 (4.0) | | 74 | 7.2 (4.0) | |
| Consumes alcohol | 487 | | | 225 | | | 262 | | |
| No | 341 | 6.5 (4.1) | **0.008**[b] | 101 | 7.4 (4.2) | 0.98[b] | 240 | 6.2 (4.1) | **0.0006**[b] |
| Yes | 146 | 7.6 (3.9) | | 124 | 7.4 (4.0) | | 22 | 8.8 (3.0) | |
| Uses sleeping pills | 487 | | | 225 | | | 262 | | |
| No | 465 | 6.9 (4.1) | 0.67[b] | 215 | 7.3 (4.0) | 0.83[b] | 250 | 6.5 (4.0) | 0.44[b] |
| Yes | 22 | 6.4 (5.0) | | 10 | 7.7 (5.0) | | 12 | 5.3 (5.0) | |
| Perceived village wealth | 485 | | | 224 | | | 261 | | |

| | | | | | | | | | |
|---|---|---|---|---|---|---|---|---|---|
| Poor or Average | 199 | 7.2 (3.6) | 0.16[b] | 99 | 8.0 (3.6) | **0.026**[b] | 100 | 6.4 (3.5) | 0.69[b] |
| Wealthy | 286 | 6.7 (4.3) | | 125 | 6.9 (4.3) | | 161 | 6.6 (4.4) | |
| Perceived village spirit | 487 | | | 225 | | | 262 | | |
| Weak or Average | 145 | 7.8 (3.8) | **0.002**[b] | 65 | 8.7 (3.4) | **0.001**[b] | 80 | 7.0 (3.9) | 0.14[b] |
| Strong | 342 | 6.5 (4.2) | | 160 | 6.9 (4.2) | | 182 | 6.2 (4.1) | |
| Cohen's Perceived Stress Scale[f] | 490 | -0.16 | **0.0003**[a] | 226 | -0.19 | **0.004**[a] | 264 | -0.11 | 0.07[a] |
| Multidimensional Scale of Perceived Social Support[f] | 490 | 0.17 | **0.0004**[a] | 226 | 0.13 | 0.06[a] | 264 | 0.21 | **0.0007**[a] |
| Perceived Social Conflict Score[f] | 490 | 0.02 | 0.63[a] | 226 | 0.11 | 0.09[a] | 264 | -0.05 | 0.46[a] |
| *Sleep measures* | *Overall (n=519)* | | | *Males (n=233)* | | | *Females (n=286)* | | |
| | n | Cor Coef or Mean (SD) | *p* | n | Cor Coef or Mean (SD) | *p* | n | Cor Coef or Mean (SD) | *p* |
| Women's Health Initiative Insomnia Rating Scale[f] | 489 | -0.08 | 0.08[a] | 225 | -0.05 | 0.45[a] | 265 | -0.10 | 0.12[a] |
| Chronotype | 477 | | | 217 | | | 260 | | |
| Morning person | 260 | 6.3 (4.2) | **0.0008**[b] | 122 | 6.5 (4.2) | **0.0005**[b] | 138 | 6.1 (4.2) | 0.12[b] |
| Evening person | 217 | 7.5 (3.8) | | 95 | 8.4 (3.6) | | 122 | 6.8 (3.9) | |
| Sleep duration (self-reported hours/night or day depending on work schedule) | 488 | -0.03 | 0.50 | 226 | -0.03 | 0.63 | 262 | -0.02 | 0.77 |
| Sleep duration categorical (self-reported hours/night or day depending on work schedule) | | | | | | | | | |
| <6 hours | 50 | 6.9 (4.1) | | 20 | 7.6 (4.1) | | 30 | 6.5 (4.2) | |
| 6-9 hours | 318 | 6.6 (4.2) | 0.20[c] | 146 | 7.2 (4.2) | 0.58[c] | 172 | 6.2 (4.2) | 0.36[c] |
| >9 hours | 121 | 7.4 (3.7) | | 60 | 7.8 (3.8) | | 61 | 7.1 (3.6) | |

Cor Coef, correlation coefficient; SD, standard deviation; AUA, Apia Urban Area; NWU, Northwest Upolu; ROU, Rest of Upolu; SF-8, short form 8; ESS includes imputed values as described in Table S2; [a]Pearson correlation; T-test; [c]Welch's one-way analysis of means (non-constant variance); [d]One-way analysis of variance (ANOVA); [e]Spearman correlation; [f]Imputation performed as described in Table S2

**Table S4.** Bivariate associations between participant characteristics and the Women's Health Initiative Insomnia Rating Scale (WHIIRS, possible scores of 0 to 20)

| *Biological factors and health history* | *Overall (n=519)* | | | *Males (n=233)* | | | *Females (n=286)* | | |
|---|---|---|---|---|---|---|---|---|---|
| | n | Cor Coef or Mean (SD) | *p* | n | Cor Coef or Mean (SD) | *p* | n | Cor Coef or Mean (SD) | *p* |
| Age (years) | 490 | -0.05 | 0.26[a] | 225 | 0.04 | 0.58[a] | 265 | -0.12 | 0.05[a] |
| Sex | 490 | | | NA | | | | | |
| Male | 225 | 6.3 (2.7) | 0.40[b] | | | | | | |
| Female | 265 | 6.6 (2.8) | | | | | | | |
| Body mass index (kg/m$^2$) | 488 | 0.18 | **0.0001**[a] | 225 | 0.13 | **0.049**[a] | 263 | 0.21 | **0.0008**[a] |
| Abdominal circumference (cm) | 488 | 0.17 | **0.0001**[a] | 225 | 0.15 | **0.029**[a] | 263 | 0.19 | **0.002**[a] |
| rs373863828 genotype | 490 | | | 225 | | | 265 | | |
| GG | 211 | 6.5 (2.6) | | 96 | 6.3 (2.6) | | 115 | 6.7 (2.7) | |
| AG | 192 | 6.3 (2.8) | 0.66[c] | 88 | 6.3 (2.8) | 0.93[c] | 104 | 6.3 (2.8) | 0.61[c] |
| AA | 87 | 6.6 (2.8) | | 41 | 6.5 (2.6) | | 46 | 6.7 (3.0) | |
| Diabetes | 490 | | | 225 | | | 265 | | |
| No | 91 | 5.9 (2.6) | | 49 | 5.6 (2.5) | | 42 | 6.3 (2.7) | |
| Pre-diabetes | 246 | 6.5 (2.7) | 0.06[c] | 111 | 6.4 (2.6) | **0.023**[c] | 135 | 6.6 (2.7) | 0.75[c] |
| Yes | 153 | 6.7 (2.9) | | 65 | 6.9 (2.7) | | 88 | 6.6 (3.0) | |
| Hypertension | 480 | | | 224 | | | 256 | | |
| No | 321 | 6.5 (2.7) | 0.76[b] | 157 | 6.4 (2.6) | 0.73[b] | 164 | 6.5 (2.7) | 0.87[b] |
| Yes | 159 | 6.4 (2.8) | | 67 | 6.3 (2.7) | | 92 | 6.5 (2.9) | |
| Asthma | 488 | | | 223 | | | 265 | | |
| No | 465 | 6.4 (2.7) | 0.44[b] | 210 | 6.3 (2.6) | 0.74[b] | 255 | 6.5 (2.8) | 0.42[b] |
| Yes | 23 | 7.0 (3.7) | | 13 | 6.7 (3.8) | | 10 | 7.5 (3.6) | |
| SF-8 Physical Health Component Score[e] | 490 | 0.06 | 0.20[a] | 225 | 0.05 | 0.46[a] | 265 | 0.07 | 0.28[a] |
| SF-8 Mental Health Component Score[e] | 490 | 0.08 | 0.08[a] | 225 | 0.09 | 0.20[a] | 265 | 0.08 | 0.22[a] |
| *Sociodemographic and behavioral factors* | *Overall (n=519)* | | | *Males (n=233)* | | | *Females (n=286)* | | |
| | n | Cor Coef or Mean (SD) | *p* | n | Cor Coef or Mean (SD) | *p* | n | Cor Coef or Mean (SD) | *p* |
| Census region | 490 | | | 225 | | | 265 | | |
| AUA | 102 | 6.4 (3.0) | | 45 | 6.1 (2.7) | | 57 | 6.6 (3.2) | |
| NWU | 211 | 6.9 (2.7) | **0.005**[c] | 101 | 6.8 (2.9) | 0.07[c] | 110 | 7.0 (2.6) | 0.06[c] |
| ROU | 177 | 6.0 (2.5) | | 79 | 5.9 (2.2) | | 98 | 6.1 (2.7) | |
| Material lifestyle score (total number of household assets)[e] | 487 | 0.07 | 0.10[a] | 224 | 0.03 | 0.67[a] | 263 | 0.12 | 0.06[a] |
| Education (years) | 489 | 0.10 | **0.024**[a] | 224 | 0.11 | 0.09[a] | 265 | 0.09 | 0.16[a] |
| Relationship status | 489 | | | 225 | | | 264 | | |
| Partnered | 398 | 6.4 (3.0) | 0.71[b] | 185 | 6.5 (2.6) | 0.18[b] | 213 | 6.5 (2.8) | 0.50[b] |
| Not partnered | 91 | 6.5 (2.7) | | 40 | 5.8 (3.0) | | 51 | 6.8 (3.0) | |
| Children at home | 489 | | | 224 | | | 265 | | |
| No | 53 | 5.9 (2.6) | 0.14[b] | 25 | 5.7 (2.5) | 0.20[b] | 28 | 6.1 (2.7) | 0.41[b] |
| Yes | 436 | 6.5 (2.7) | | 199 | 6.4 (2.7) | | 237 | 6.6 (2.8) | |
| Physical activity | 488 | | | 223 | | | 265 | | |
| 0 MVPA minutes/day | 360 | 6.5 (2.8) | 0.21[b] | 138 | 6.5 (2.7) | 0.41[b] | 222 | 6.6 (2.8) | 0.50[b] |
| >0 MVPA minutes/day | 128 | 6.2 (2.7) | | 85 | 6.2 (2.6) | | 43 | 6.3 (2.9) | |
| Work schedule | 489 | | | 224 | | | 265 | | |
| Day shift | 226 | 6.7 (2.6) | | 115 | 6.7 (2.4) | | 111 | 6.8 (2.9) | |
| Afternoon or night shift | 22 | 6.4 (3.2) | **0.0003**[c] | 11 | 6.6 (2.9) | **0.003**[c] | 11 | 6.1 (3.6) | **0.042**[c] |
| Other [split, irregular/on-call, rotating] | 51 | 4.9 (2.8) | | 18 | 4.2 (2.4) | | 33 | 5.3 (2.9) | |
| Does not work | 190 | 6.5 (2.7) | | 80 | 6.3 (2.9) | | 110 | 6.7 (2.5) | |
| Owns or rents home | 487 | | | 223 | | | 264 | | |
| Owns | 463 | 6.5 (2.7) | 0.11[b] | 216 | 6.3 (2.6) | 0.88[b] | 247 | 6.6 (2.8) | 0.07[b] |
| Rents | 24 | 5.6 (2.6) | | 7 | 6.1 (2.7) | | 17 | 5.4 (2.6) | |
| Smokes | 489 | | | 225 | | | 264 | | |
| No | 298 | 6.5 (2.7) | 0.92[b] | 108 | 6.5 (2.6) | 0.39[b] | 190 | 6.5 (2.8) | 0.32[b] |
| Yes | 191 | 6.5 (2.8) | | 117 | 6.2 (2.7) | | 74 | 6.8 (2.9) | |
| Consumes alcohol | 487 | | | 224 | | | 263 | | |
| No | 341 | 6.5 (2.8) | 0.98[b] | 100 | 6.4 (2.7) | 0.91[b] | 241 | 6.5 (2.8) | 0.29[b] |
| Yes | 146 | 6.5 (2.6) | | 124 | 6.3 (2.6) | | 22 | 7.1 (2.6) | |
| Uses sleeping pills | 487 | | | 224 | | | 263 | | |
| No | 465 | 6.5 (2.7) | 0.09[b] | 214 | 6.4 (2.7) | 0.20[b] | 251 | 6.6 (2.8) | 0.31[b] |
| Yes | 22 | 5.5 (2.6) | | 10 | 5.2 (2.7) | | 12 | 5.8 (2.6) | |

| | | | | | | | | | |
|---|---|---|---|---|---|---|---|---|---|
| Perceived village wealth | 485 | | | 223 | | | 262 | | |
| Poor or Average | 200 | 5.8 (2.7) | **1.02E-05**[b] | 99 | 5.9 (2.8) | **0.013**[b] | 101 | 5.8 (2.6) | **0.0003** [b] |
| Wealthy | 285 | 6.9 (2.7) | | 124 | 6.7 (2.5) | | 161 | 7.1 (2.8) | |
| Perceived village spirit | 487 | | | 224 | | | 263 | | |
| Weak or Average | 145 | 5.8 (2.7) | **0.001**[b] | 64 | 6.0 (2.7) | 0.17[b] | 81 | 5.8 (2.8) | **0.001**[b] |
| Strong | 342 | 6.7 (2.7) | | 160 | 6.5 (2.6) | | 182 | 6.9 (2.8) | |
| Cohen's Perceived Stress Scale[e] | 490 | -0.01 | 0.83[a] | 225 | -0.06 | 0.38[a] | 265 | 0.03 | 0.65[a] |
| Multidimensional Scale of Perceived Social Support[e] | 490 | 0.07 | 0.11[a] | 225 | 0.15 | **0.023**[a] | 265 | 0.03 | 0.65[a] |
| Perceived Social Conflict Score[e] | 490 | 0.05 | 0.26[a] | 225 | -0.03 | 0.70[a] | 265 | 0.11 | 0.07[a] |
| *Sleep measures* | *Overall (n=519)* | | | *Males (n=233)* | | | *Females (n=286)* | | |
| | n | Cor Coef or Mean (SD) | *p* | n | Cor Coef or Mean (SD) | *p* | n | Cor Coef or Mean (SD) | *p* |
| Chronotype | 477 | | | 216 | | | 261 | | |
| Morning person | 260 | 6.6 (2.8) | 0.25[b] | 121 | 6.6 (2.6) | 0.10[b] | 139 | 6.6 (2.9) | 0.88[b] |
| Evening person | 217 | 6.3 (2.6) | | 95 | 6.1 (2.6) | | 122 | 6.6 (2.7) | |
| Sleep duration (self-reported hours/night or day depending on work schedule) | 488 | 0.01 | 0.76 | 225 | 0.06 | 0.40 | 263 | -0.3 | 0.69 |
| Sleep duration categorical (self-reported hours/night or day depending on work schedule) | | | | | | | | | |
| <6 hours | 50 | 5.9 (2.7) | 0.08[c] | 20 | 5.8 (1.9) | **0.016**[c] | 30 | 6.0 (3.1) | 0.50[c] |
| 6-9 hours | 318 | 6.7 (2.6) | | 146 | 6.7 (2.5) | | 172 | 6.6 (2.7) | |
| >9 hours | 121 | 6.2 (3.1) | | 60 | 5.6 (3.1) | | 61 | 6.7 (2.9) | |

Cor Coef, correlation coefficient; SD, standard deviation; AUA, Apia Urban Area; NWU, Northwest Upolu; ROU, Rest of Upolu; SF-8, short form 8; WHIIRS includes imputed values as described in Table S2; [a]Pearson correlation; T-test; [c]One-way analysis of variance (ANOVA); [d]Spearman correlation; [e]Imputation performed as described in Table S2

**Figure S1.** Scatterplot depicting correlation between Women's the Health Initiative Insomnia Rating Scale (WHIIRS) and the Epworth Sleepiness Scale (ESS)

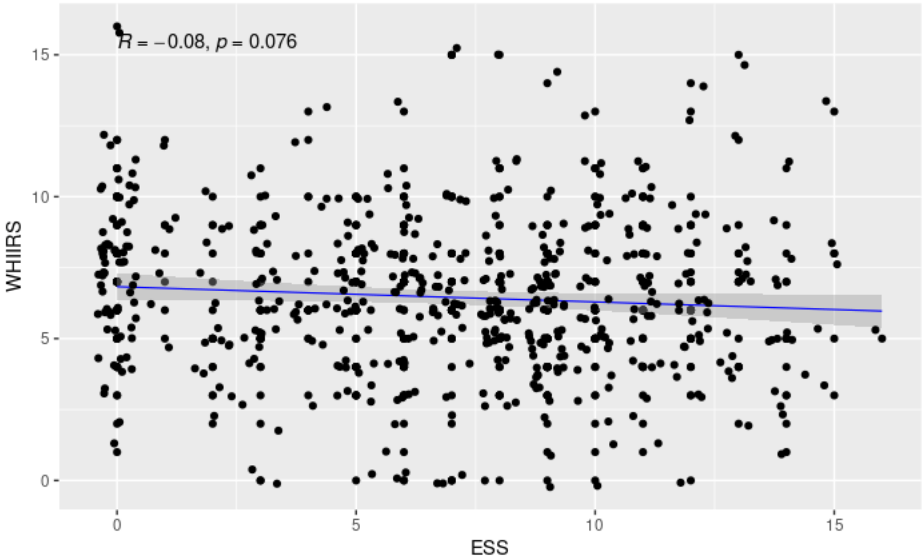


WHIIRS, Women's Health Initiative Insomnia Rating Scale; ESS, Epworth Sleepiness Scale; R, Pearson's correlation coefficient; blue solid line, fitted regression line
